# Evaluating User Experiences with an AI Chatbot for Health-Related Social Needs: A Cross-Sectional Mixed Methods Study

**DOI:** 10.1101/2025.06.16.25329054

**Authors:** Emre Sezgin, Daniel I. Jackson, Syed-Amad Hussain, A. Baki Kocaballi, Carson Richardson, Micah Skeens, Eric Fosler-Lussier, Aldenise P. Ewing, Macarius Donneyong, Ahna Pai

## Abstract

**Background:** Unmet Health-related social needs (HRSNs), such as food insecurity and housing, significantly impact health outcomes and wellbeing. Although screening tools are widely adopted to identify the needs, sustainable linkage to resources remains challenging. Conversational agents (chatbots) offer potential solutions for tailored and personalized feedback, real-time navigation, yet their usability and trustworthiness among populations with high needs require further exploration.

**Methods:** We conducted a mixed-methods study to evaluate user experiences with the DAPHNE© chatbot, which is designed to identify unmet HRSNs and provide personalized resource recommendations. Quantitative and qualitative data were collected from 128 caregivers with at least one dependent child. Online study design combined scenario/ task-based and free form chatbot use, to guide the engagement. Study measures included usability (SUS), task load (NASA-TLX), satisfaction (NPS), and trust. Qualitative analysis involved user feedback and user-chatbot conversation transcripts. We used regression and pairwise analyses to explore associations between demographic characteristics, self-reported unmet HRSNs and user experience outcomes (usability, satisfaction, task load and trust).

**Results:** Most participants were female (68%), aged 30-49 years (71%), and White (44%) or Black/African American (36%) and Hispanic/Latino (27%), relied on Medicaid/Medicare (83%), and cared for a child with, or had, special healthcare needs (78%). Participants reported high usability (SUS= 84.7, SD=12.4), low task load (NASA-TLX= 6.8, SD=2.8), high satisfaction (NPS= 8.0, SD=2.4), and high trust (Mean= 4.1, SD=0.8). Nearly all participants (98%) reported unmet HRSNs, including food insecurity (76%) and financial limitations (75%). Free-form chatbot conversation sessions averaged 3 minutes and ∼20 turns, with greater use of assistive buttons over typing. Furthermore, DAPHNE (using retrieval-augmented generation to ground every recommendation in a live social-care API) achieved 99 % intent accuracy in 1,523 message turns. Dialogues focused on financial, housing, and food needs. 94% of participants found the tool was helpful, while requesting design features like saved histories, voice interaction, and richer local resource details. Regression analysis showed usability and trust were broadly consistent across most demographic groups, though participants with higher education and lower income showed modest decrements in usability. Several HRSNs, including transportation and utility disruptions, were associated with higher trust and satisfaction, suggesting the assistant may hold particular value for users facing structural these barriers.

**Discussion:** The DAPHNE chatbot demonstrates potential as a useful tool for addressing HRSNs, with strong usability and trust among diverse populations. Future designs should focus on longitudinal impact assessments and effectiveness to enhance accessibility and address practical implementation challenges.

## Introduction

Health-related social needs (HRSNs), such as housing stability, transportation, food security, access to healthcare, and financial stability, account for over half of modifiable health outcomes and are closely associated with elevated morbidity, increased healthcare utilization, and premature mortality.^1–3^ These needs cluster along entrenched lines of income, race, and geography, amplifying inequities in health outcomes.^4,5^ Children are particularly vulnerable: persistent material deprivation impairs physical, emotional, and cognitive development and predicts poorer adult health outcomes.^4–7^ Routine HRSN screening is recommended across pediatric and adult healthcare settings, prompting widespread adoption of brief screeners in primary care, emergency departments, and inpatient units.^7^ However, screening alone is insufficient; linking families to community resources remains a critical challenge.^8,9^

In many clinics, a positive screening yields only brief provider guidance and a printed “resource sheet,” leaving patients and caregivers to navigate a fragmented constellation of supports. Staffing constraints, language discordance, and shifting eligibility requirements often derail this passive model - fewer than half of referred patients ultimately receive services.^10,11^ While electronic health record-embedded screeners and patient portals enhance data capture, their effectiveness is limited without integration into a dynamic referral system. Their reliance on static directories and manual matching heightens the risk of outdated information and missed connection..^12–14^ Conversational agents (chatbots) can bridge the gap in understanding needs and navigating resources by coupling natural-language processing with adaptive dialogue to deliver tailored guidance at scale.^15,16^ Across the literature of chatbot use in healthcare, systematic reviews and pilot studies focusing on health-information seeking, preventive screening, medication adherence, and chronic-disease self-management consistently report high perceived usability and promising clinical impact.^17–21^

Our prior iterative design and evaluation study of a semi-functional HRSN chatbot, demonstrated perceived utility for resource linkage but also highlighted concerns regarding accuracy, resource sharing, and user trust among adult caregivers.^18^ The current study builds upon this earlier work with an improved and fully functional DAPHNE (Dialog-based Assistant Platform for Healthcare and Needs Ecosystem) chatbot.^18^ Specifically, this study aimed to evaluate the user experience associated with the DAPHNE chatbot among caregivers with unmet HRSNs. We sought to understand how users engage with an AI-powered social care assistant and to identify demographic or contextual factors associated with varied user experiences. The findings of this study will inform future design enhancements and implementation strategies for equitably scaling digital social care interventions across diverse clinical and community settings.

## Methods

### Study Design

We conducted a mixed-methods study with a convergent parallel design to evaluate DAPHNE chatbot.^22^ Specifically, we assessed user experience with measures of usability, satisfaction, task load, and trust, while examining how sociodemographic factors and identified HRSNs moderate these outcomes. The reporting of this mixed-methods observational study was guided by the STROBE checklist, ensuring transparency and completeness in the presentation of methods and results. **Figure 1** outlines the overall study design and procedures.

**Figure 1.**
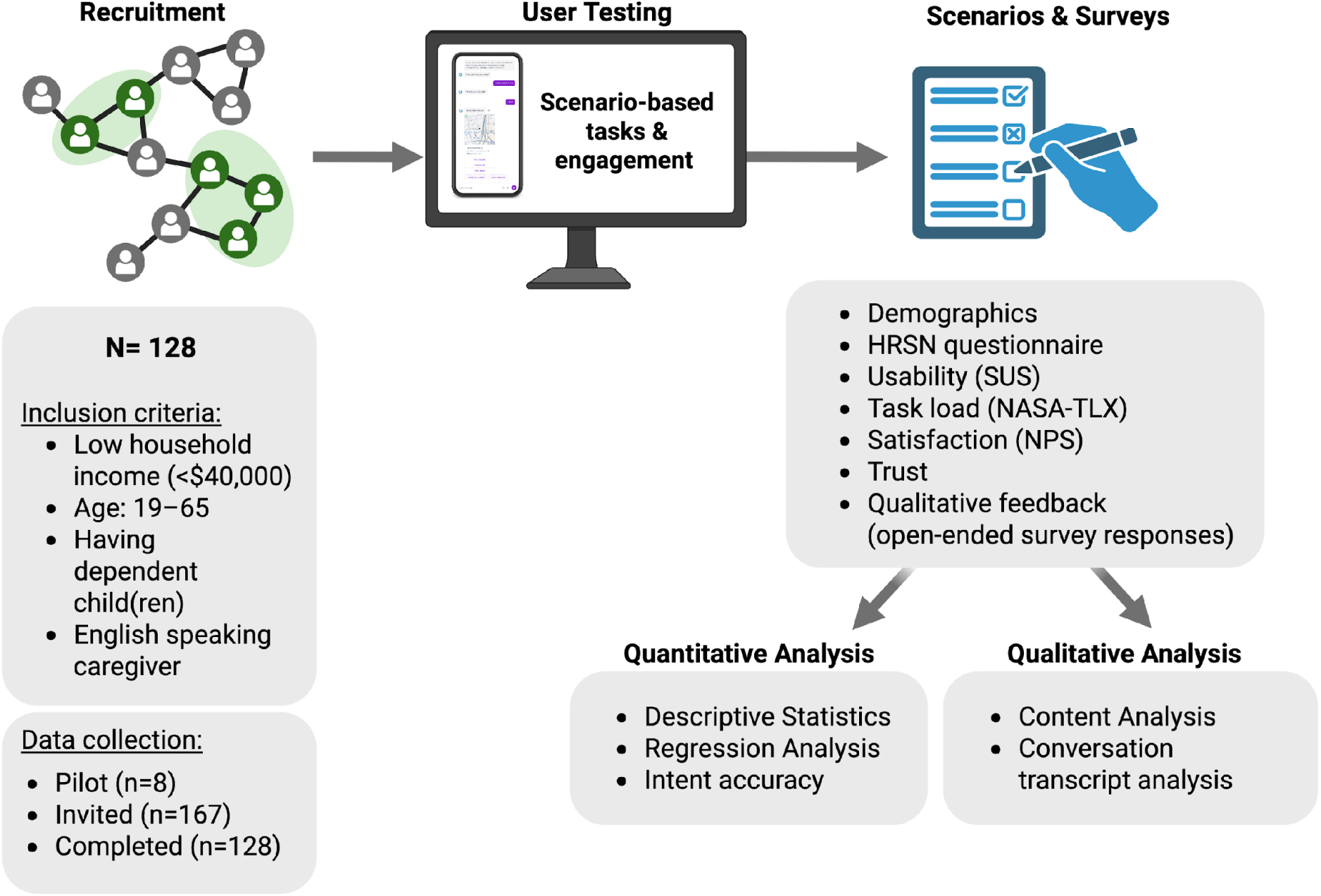
Study design, procedure and data collection.

### DAPHNE Chatbot

The DAPHNE chatbot was developed to identify unmet HRSNs and provide personalized, location-based community resource recommendations via preferred natural language. The chatbot employed a semi-structured conversational interface that supported both free-text and button-based inputs, allowing flexible user interaction and navigating community resources. After completing each session, participants were prompted to rate their experience and provide feedback, including both the quality of resource information and the conversational flow.

To design and prototype the dialog system, Amazon Web Services (AWS) was initially used to test conversation flows and explore integration of early large language model (LLM) capabilities, including intent recognition and generation of empathetic responses. Once core functions were finalized, the chatbot was adapted to a web app (accessible via desktop or mobile device web browser) using Voiceflow platform that supports secure, browser-based deployment and dialog management, including integration with third-party APIs and LLMs for natural language tasks. **Figure 2** provides an outline of DAPHNE chatbot and its ecosystem.

**Figure 2.**
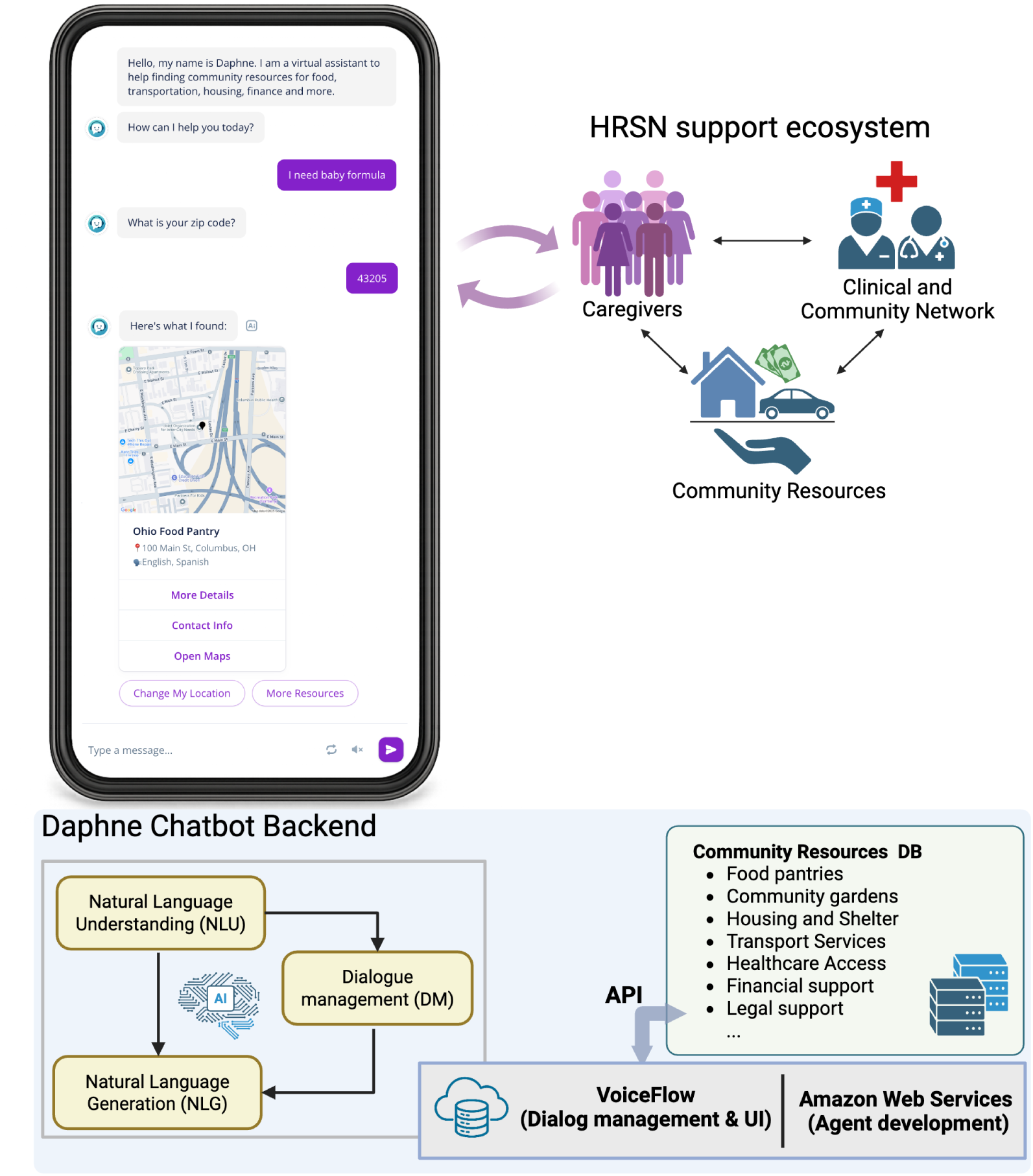
Overview of the DAPHNE chatbot. This system uses Natural Language Understanding and Application Programming Interface for its functionality and to augment existing HRSN support

To accommodate variability in user interaction styles (such as button selection versus typed input) the chatbot used a hybrid dialogue architecture that combined rule-based logic for structured flows (e.g., asking for zip code, presenting resource categories) with Natural Language Understanding (NLU) for more open-ended queries. For example, if a user typed, “I need help paying rent,” the NLU module mapped this intent to the “housing support” domain and triggered the corresponding response logic. NLU and Natural Language Generation (NLG) were powered by large language models (OpenAI’s GPT-3.5 Turbo and Anthropic’s Claude 3.5 Haiku),^23,24^ which helped interpret ambiguous input, clarify misunderstood queries, and provide empathetic language in follow-up prompts. In cases where the chatbot could not confidently categorize a user message-for example, if the input did not match any predefined intent or the user entered a vague or unrelated phrase-this was flagged as a non-categorized entry, triggering a fallback script. The fallback response informed users that the chatbot could not understand their request and prompted them to try again or use button options.

Recommendations were generated based on data retrieved from a live social-care API (Findhelp.org), which contained up-to-date community resource listings. The chatbot only used information returned from the API and did not generate recommendations independently, ensuring response accuracy and grounding in real-world availability. Some user requests, such as highly specific eligibility questions, were occasionally not available through the API, and in such cases, users were informed accordingly. This structured query-and-response design follows the Retrieval-Augmented Generation (RAG) framework provided by the platform, which combines real-time information retrieval with natural language output to ensure personalized and contextually relevant guidance.^25,26^

### Recruitment and procedure

Participants were recruited through an online research platform (Prolific.com) following a purposeful sampling approach. Recruitment and study participation occurred between January 1st and March 30th, 2025. Inclusion criteria required participants to be caregivers aged 19 years or older, residing in the U. S., caring for at least one dependent child, and reporting an annual household <$40,000 (<200% of the 2025 United States Federal Poverty Level). Participants were excluded from the study if they declined consent, opted out at any step of the study, or failed attention-check tasks embedded within chatbot interactions. Informed consent was obtained electronically, and participants completed study participation via a HIPAA-compliant online survey platform (REDCap).^27^ Participants were compensated for their time ($12). Recruitment ended after exhausting funding for compensation.

Participants received their instructions and completed quantitative and qualitative measures on REDCap before and after engaging with the chatbot. Engagement with the chatbot was guided through four scenarios. Prior to the data collection, a pilot group of participants (N=8) were recruited to test for understandability and clarity of survey questions, scenarios and chatbot conversations. In response to their feedback, we revised the study instructions in the survey, added clarifications to HRSN-related questions, and refined scenario content to improve understandability. The data from pilot participants were not included in the analysis.

#### Scenarios

The study team designed four scenarios, informed by our previous investigation,^18^ to simulate real-world cases and to demonstrate the chatbot’s capabilities. Scenarios were used to standardize the content that the participants engaged with (details in **Supplementary Appendix 1**). The scenario topics were selected purposefully to reflect high-priority HRSNs - food insecurity, healthcare access, and housing instability - that align with Center for Medicare and Medicaid Services (CMS) screening requirements and commonly targeted domains in Medicaid and health system initiatives.^28^

In addition, each scenario tested a distinct chatbot function (e.g., location-based search, cost inquiry, service coverage), allowing structured evaluation of key features. The final open-ended task enabled observation of natural user behavior, offering a balanced assessment of performance and real-world engagement. Specifically, the first scenario required participants to identify food and nutritional assistance programs and obtain information about the operational hours of these programs based on their residential zip codes. In the second scenario, participants explored healthcare-related resources in a new zip code and requested information regarding the new program’s cost of service. The third scenario involved participants seeking housing support resources in a nearby city, focusing specifically on determining service coverage and obtaining relevant contact information. Following the completion of these structured scenarios, the last scenario instructed participants to freely engage and explore the chatbot’s capabilities in line with their personal interests or social needs.

#### Data Collection

Quantitative data collection included demographic characteristics, HRSNs, and user experience. The HRSNs assessed in the survey comprised food insecurity, housing instability, transportation barriers, financial strain, social isolation, and health literacy deficits. Items for measuring HRSNs were adapted from established screening instruments utilized by the U.S. Census Bureau,^29^ advocacy organizations,^30–32^ healthcare systems,^33,34^ research institutes,^35–39^ and community health centers. User experience was assessed using four standardized instruments: the National Aeronautics Space Administration Task Load Index (NASA-TLX),^40^ System Usability Scale (SUS),^41^ Net Promoter Score (NPS),^42^ and a Trust scale (adapted from Jian et al.).^43^

Qualitative data were collected via open-ended responses within REDCap. Specifically, participants provided narrative responses to describe a positive experience, a negative experience, and suggested improvements for the chatbot. In addition, transcripts of participant-hatbot interactions were recorded.

### Data Analysis

We reported descriptives of demographics, HRSN and user experience responses. In addition, we conducted qualitative analysis of open-ended survey responses, conversational analysis using transcripts of chatbot-user engagement, calculated chatbot intent accuracy and finally exploratory analysis to investigate the association between demographics, HRSN vs. user experience outcomes via univariable regression models.

#### Qualitative analysis

Qualitative data were two-fold: (1) Free-text feedback over the survey and (2) chatbot conversation transcripts. Free-text feedback was analyzed for sentiment (i.e., positive, negative, and neutral) and intensity (i.e., high, low) to highlight interactions with the chatbot. Sentiment was rated by the study team (AH, DIJ) on a Likert scale from 1 (Negative) to 5 (Positive) indicating the level of satisfaction in the user’s language. Intensity similarly coded by the study team on a Likert scale from 1 (Low) to 5 (High) indicating the level of emotional content in user’s language (see Supplementary Appendix 2). Furthermore, we conducted a thematic analysis, developing an initial codebook through an inductive review of 10% of the transcripts (DJ, AH). Codes were refined iteratively after each transcript was reviewed. After the final transcript, codes were reviewed once more to define our themes. Organization of transcription data was processed through Python,^44^ and coding of the dataset was done through Microsoft Excel. Interrater reliability was assessed using Cohen’s Kappa.^45^ Any disputes were resolved by a senior team member (ES).

#### Conversational analysis

Chatbot conversation transcripts were coded and manually analyzed by research team members (DIJ, AH) using a structured rubric to categorize patterns in resource-seeking behaviors for each free-form conversational turn. The defined rubric notes when users mention (1) a specific resource, (2) the purpose for the resource, (3) their personal background, (4) multiple needs, and (5) thoughts on finding a resource (see Supplementary Appendix 3). Each behavior was a binary measure where present strategies were labeled as 1 and absent strategies were labeled as 0. A *specific resource* included a title or descriptor to explain their need (for example, “allergen-free food”). A *purpose for the resource* included mentions on who or why the resource is necessary (“for my friend”). The *personal background* referred to contextual details about their circumstances (“I just moved”). *Multiple needs as a resource-seeking* behavior including at least two distinct needs that would require different types of assistance (such as across multiple HRSNs). Finally, we had the category for participants expressing *their thoughts on finding a resource* with implicit or explicit intent of exploring community resources.

#### Intent accuracy

To validate the chatbot’s NLU accuracy (ability to recognize HRSNs from the user’s free-form entry), the study team calculated user “intents” as a percentage of accuracy. For each message that requires a fallback mechanism from the chatbot to correctly identify the topic of interest, the intent is approximated out of the possible services available in the API. We automatically tagged each occurrence in the chatbot’s infrastructure while exporting transcription data.

#### Regression analysis

Chatbot usability, satisfaction, and task load were modeled via linear regression, whereas chatbot trustworthiness was modeled using an ordinal logistic regression. More specifically, we performed category-wise regressions where the coefficients of each variable in the category are compared against the category’s reference variable (RV).

For each demographic category, regression coefficients were interpreted relative to a reference variable (RV), which is displayed as the first category in **Figure 3**. Demographic variables included both binary (e.g., Insurance: Yes/No) and categorical predictors with multiple response levels (e.g., Marital Status). For multi-level categorical predictors, we highlight response options with “statistically significant” coefficients (i.e., 95% confidence intervals that do not cross the null value defined by the RV). Where appropriate, we also present binarized contrasts (e.g., Married vs. all other marital statuses). Furthermore, for trust factor, we presented predicted mean trust level to directly correlate level of trust to each demographic and HRSN group in **Supplementary Appendix S10.1 - S10.7**.

**Figure 3.**
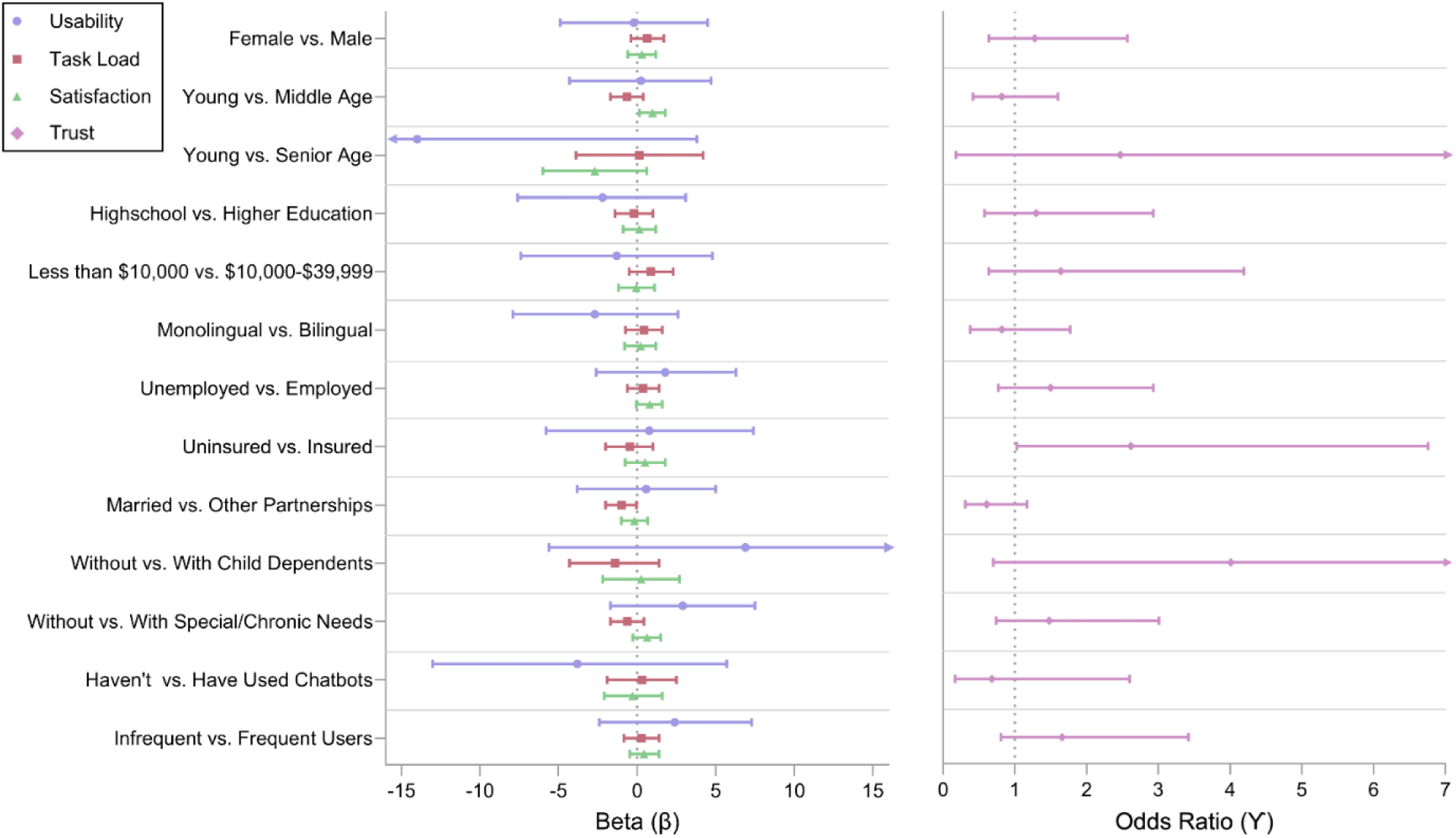
Regression analysis forest plot for demographics (binary) and user experience. First variable in each category is the reference value.

We additionally conducted a pairwise analysis between variables within a given category (e.g. social isolation) to quantify suggestive trends between user demographics and HRSN against our user experience metrics. Pairwise results were noted in terms of differences between variable means (Beta difference for Usability, Task Load, and Satisfaction) and are presented in **Supplementary Appendix S9.1-S9.20**. Suggestive trends were presented either as monotonically scaling, especially for ordinal variables (e.g. education or income), or general shifts. All statistical analyses were conducted using statistical software, R (v4.5.0).

## Results

We recruited 167 participants to the study, and 128 participants (76.6%) completed all activities and were included in the analysis.

### Demographics

Participants were geographically distributed across diverse urban and rural settings throughout the U.S. (**Table 1**). The sample (N=128) was predominantly White (44%) and Black/African American (36%), with 27% identifying as Hispanic or Latino. Most participants identified as female (68%), were between 30 and 49 years old (71%), and primarily spoke only English (78%). Although 22% were bilingual, the most spoken second language was Spanish (64% of bilingual participants). Participants typically had varied educational backgrounds, with the majority having completed some college or an associate/bachelor’s degree (69%). Most were employed (60%), and reported an annual household income between $20,000 and $39,999 (73%). A significant majority relied on Medicaid or Medicare insurance (83%). Participants were commonly married or in an unmarried partnership (54%), while about one-third (34%) reported never being married. Nearly all participants cared for at least one dependent child (97%), or had 1t o 2 children (78%), most frequently aged between 6 and 12 years old (37%). Most participants (78%) reported themselves or their dependent having special healthcare needs. Participants also reported prior experience with chatbots (95%), including voice assistants like Siri and Alexa.

**Table 1.**
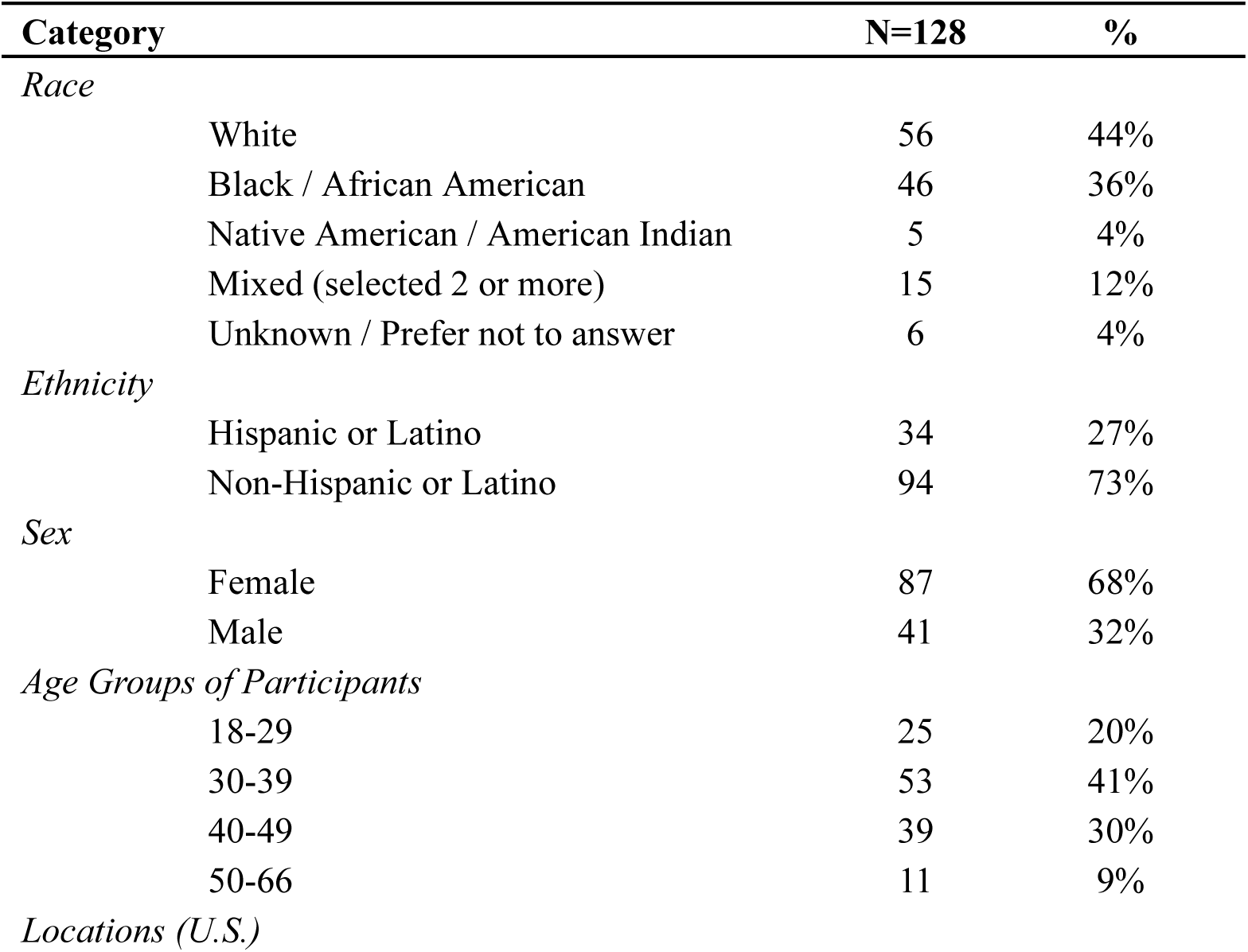
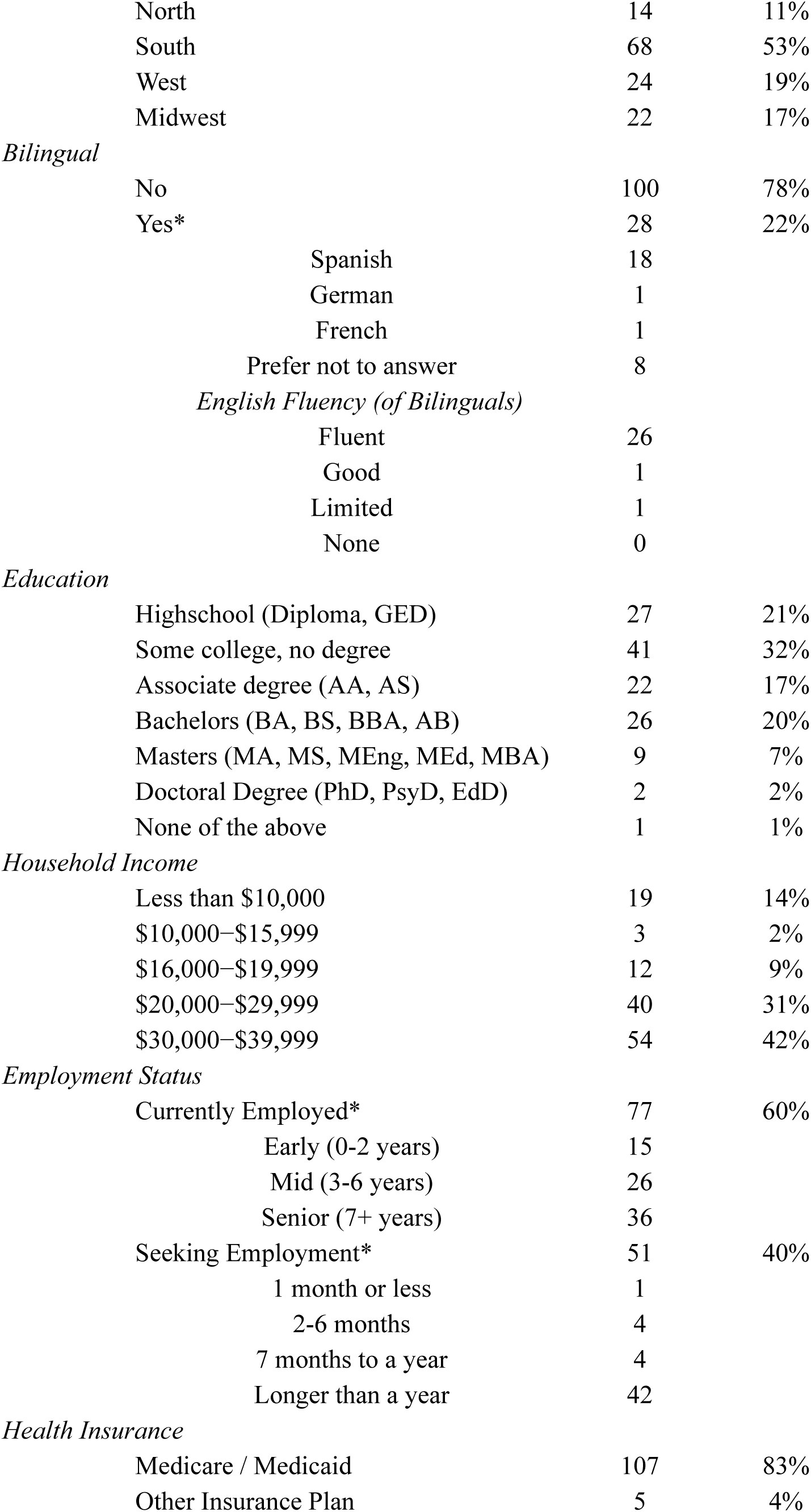
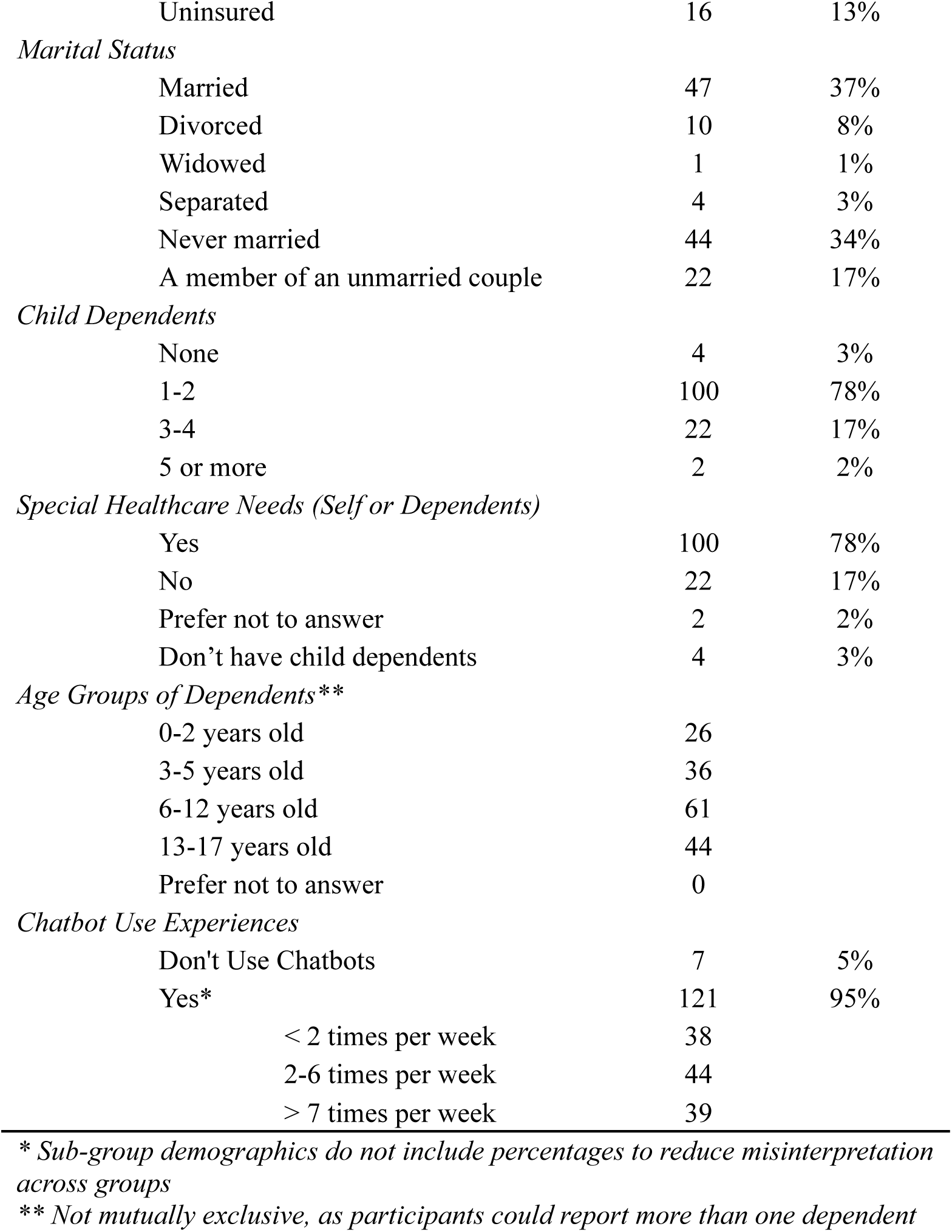
Demographic Characteristics of Participants.

| Category | N=128 | % |
| --- | --- | --- |
| *Race* |  |  |
| White | 56 | 44% |
| Black / African American | 46 | 36% |
| Native American / American Indian | 5 | 4% |
| Mixed (selected 2 or more) | 15 | 12% |
| Unknown / Prefer not to answer | 6 | 4% |
| *Ethnicity* |  |  |
| Hispanic or Latino | 34 | 27% |
| Non-Hispanic or Latino | 94 | 73% |
| *Sex* |  |  |
| Female | 87 | 68% |
| Male | 41 | 32% |
| *Age Groups of Participants* |  |  |
| 18-29 | 25 | 20% |
| 30-39 | 53 | 41% |
| 40-49 | 39 | 30% |
| 50-66 | 11 | 9% |
| *Locations (U.S.)* |  |  |

|  |  |  |  |
| --- | --- | --- | --- |
|  | North | 14 | 11% |
|  | South | 68 | 53% |
|  | West | 24 | 19% |
|  | Midwest | 22 | 17% |
| *Bilingual* |  |  |  |
|  | No | 100 | 78% |
|  | Yes* | 28 | 22% |
|  | Spanish | 18 |  |
|  | German | 1 |  |
|  | French | 1 |  |
|  | Prefer not to answer | 8 |  |
|  | *English Fluency (of Bilinguals)* |  |  |
|  | Fluent | 26 |  |
|  | Good | 1 |  |
|  | Limited | 1 |  |
|  | None | 0 |  |
| *Education* |  |  |  |
|  | Highschool (Diploma, GED) | 27 | 21% |
|  | Some college, no degree | 41 | 32% |
|  | Associate degree (AA, AS) | 22 | 17% |
|  | Bachelors (BA, BS, BBA, AB) | 26 | 20% |
|  | Masters (MA, MS, MEng, MEd, MBA) | 9 | 7% |
|  | Doctoral Degree (PhD, PsyD, EdD) | 2 | 2% |
|  | None of the above | 1 | 1% |
| *Household Income* |  |  |  |
| | Less than \$10,000 | 19 | 14% |
| | \$10,000–\$15,999 | 3 | 2% |
| | \$16,000–\$19,999 | 12 | 9% |
| | \$20,000–\$29,999 | 40 | 31% |
| | \$30,000–\$39,999 | 54 | 42% |
| *Employment Status* |  |  |  |
|  | Currently Employed* | 77 | 60% |
|  | Early (0-2 years) | 15 |  |
|  | Mid (3-6 years) | 26 |  |
|  | Senior (7+ years) | 36 |  |
|  | Seeking Employment* | 51 | 40% |
|  | 1 month or less | 1 |  |
|  | 2-6 months | 4 |  |
|  | 7 months to a year | 4 |  |
|  | Longer than a year | 42 |  |
| *Health Insurance* |  |  |  |
|  | Medicare / Medicaid | 107 | 83% |
|  | Other Insurance Plan | 5 | 4% |
| Uninsured | 16 | 13% |
| *Marital Status* |  |  |
| Married | 47 | 37% |
| Divorced | 10 | 8% |
| Widowed | 1 | 1% |
| Separated | 4 | 3% |
| Never married | 44 | 34% |
| A member of an unmarried couple | 22 | 17% |
| *Child Dependents* |  |  |
| None | 4 | 3% |
| 1-2 | 100 | 78% |
| 3-4 | 22 | 17% |
| 5 or more | 2 | 2% |
| *Special Healthcare Needs (Self or Dependents)* |  |  |
| Yes | 100 | 78% |
| No | 22 | 17% |
| Prefer not to answer | 2 | 2% |
| Don't have child dependents | 4 | 3% |
| *Age Groups of Dependents\*\** |  |  |
| 0-2 years old | 26 |  |
| 3-5 years old | 36 |  |
| 6-12 years old | 61 |  |
| 13-17 years old | 44 |  |
| Prefer not to answer | 0 |  |
| *Chatbot Use Experiences* |  |  |
| Don't Use Chatbots | 7 | 5% |
| Yes* | 121 | 95% |
| < 2 times per week | 38 |  |
| 2-6 times per week | 44 |  |
| > 7 times per week | 39 |  |

\* Sub-group demographics do not include percentages to reduce misinterpretation across groups
\*\* Not mutually exclusive, as participants could report more than one dependent

### Health-related Social Needs

Nearly all participants who engaged with the chatbot reported at least one unmet HRSN (98%). In Table 2, we provided combined scores (binary; y/n) of HRSNs grouped by short-term needs, long-term needs, and time-independent needs (full responses are available in Supplementary Appendix 4). The most frequently reported short-term (within the past 30 days) needs included food insecurity (75%), financial limitations (75%), and transportation barriers (57%). Food insecurity (71%) was also the most frequently reported long-term (within the past 12 months) need, followed by transportation barriers (48%) and financial limitations (38%). Additionally, while relatively few participants reported recent physical isolation from social activities (11%), a substantial number reported persistent feelings of social isolation (65%) without a specified time frame. Smaller proportions indicated difficulties related to overall literacy (reading and comprehension; 10%) and medical literacy confidence in managing health-related issues (9%).

**Table 2.** Frequencies of user-reported health-related social needs.

| HRSNs | N= 128 | % |
| --- | --- | --- |
| *Short Term (30d need)* |  |  |
| Housing | 59 | 46% |
| Food | 97 | 75% |
| Transport | 73 | 57% |
| Finance | 96 | 75% |
| Isolation | 14 | 11% |
| *Long Term (12m need)* |  |  |
| Housing | 36 | 28% |
| Food | 91 | 71% |
| Transport | 62 | 48% |
| Finance | 49 | 38% |
| *Time-Independent* |  |  |
| Isolation | 83 | 65% |
| Literacy (overall) | 13 | 10% |
| Literacy (medical) | 12 | 9% |
| *No HRSNs disclosed* | 2 | 2% |

### User Experience

As reported in Table 3, participants rated the chatbot as being highly usable (mean = 84.7, SD = 12.4), trustworthy (mean = 4.1, SD = 0.8), and satisfactory (mean = 8.0, SD = 2.4). The chatbot was rated around the lowest third of the task load index (mean = 6.8, SD = 2.8).

**Table 3.**
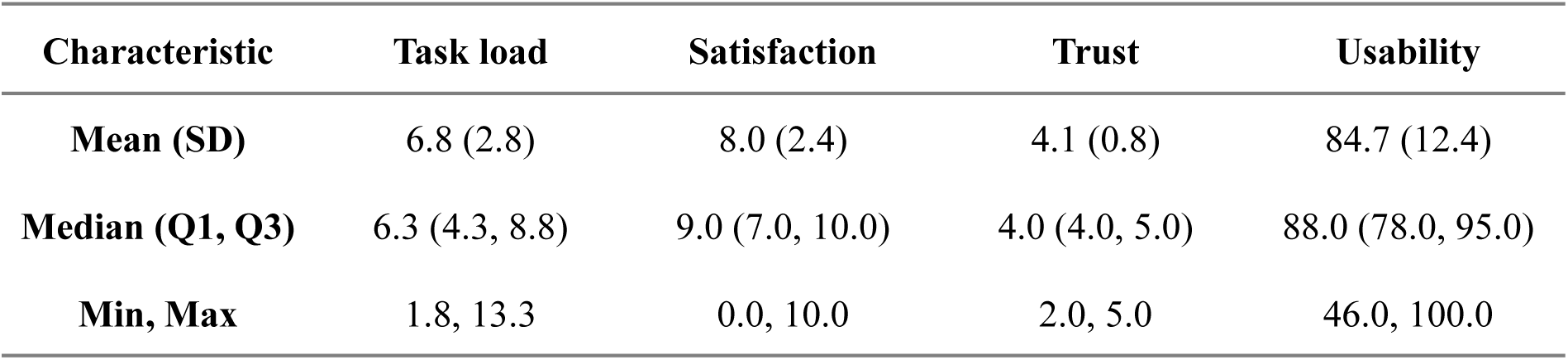
Descriptive results on user experience categories.

### Exploratory analysis

We examined the association between demographic factors and user experience metrics using category-wise regression models.

#### Demographic Factors and User Experience

No statistically significant associations were observed between usability and trust ratings vs. demographic variables (**Figure 3**). Perceived burden (Task Load) was significantly lower among participants who were Divorced (β = –2.7; 95% CI: –4.6, –0.85), Never Married (β = –1.3; 95% CI: –2.4, –0.14), and Unmarried (β = –1.4; 95% CI: –2.8, 0.00), relative to those who were Married. When aggregated, all non-married statuses were associated with significantly lower Task Load scores (β = –1.5; 95% CI: –2.5, –0.46). Satisfaction ratings were significantly higher among middle-aged participants compared to young adults (β = 0.99; 95% CI: 0.14, 1.8). See **Figure 3** for forest plots and Supplementary Appendix 4, Table S4.2 for regression results.

In terms of monotonic trends, most of the metrics scaled up with income from $10k onwards. Specifically, the most pronounced pairwise contrast (($30k-$40k) - ($10k-$16k)) showed an estimated difference between the mean of Usability (Estimate= 8.85, SE= 7.40), Task Load (Estimate= 1.28, SE= 1.69), and Satisfaction (Estimate= 3.39, SE= 1.41), with differences scaling down as income decreased. It should be noted, incomes below $10k do not follow this trend. Likewise, a pronounced estimated difference was observed between ‘Masters degree’ - ‘Some college, no degree’ in terms of task load (Estimate= 3.17, SE= 1.00), with task load estimated difference scaling down as education scales down. Education at the high school level, however, did not follow this trend as it only scaled down to ‘Some college, no degree’. Suggestive trends are presented in Supplementary Appendix (Forest plots: Figures S8.1-S8.4; pairwise metrics: Tables S9.1-S9.9, trust metrics: S10.1-S10.3).

Meanwhile, in terms of relationships which are not monotonically scaling but suggest a general shift, having income >=$16k correlates with increased Trust (Mean class of 2.97-3.12 compared to the mean class of 2.90 for incomes <$10k)). Likewise, having any post-secondary degree trends negatively for usability relative to the RV (Range of difference in estimated means = −3.72 to −5.91)). We additionally see that compared to the RV (No Children), usability (Range of difference in estimated means = 6.58 to 16.00)) trust (Mean class of 3.08 to 3.44 compared to 2.49 for no children), and satisfaction (Range of difference in estimated means = 0.14 to 2.25) trend positively with having any children while task load (Range of difference in estimated means = −1.33 to −1.7) correlates negatively.

#### Health Related Social Needs and User Experience

We examined associations between HRSNs and user experience metrics (see **Figure 4** for forest plots; regression results are available in **Supplementary Appendix 4**, **Table 3**). No statistically significant associations were observed between HRSNs and usability. Participants who reported that transportation issues prevented them from attending medical appointments had significantly higher trust ratings compared to those without such barriers (OR = 3.21; 95% CI: 1.03, 10.7). Users who reported being “very often worried about food” reported significantly lower task load scores than those who were never worried (β = –1.8; 95% CI: –3.6, –0.11). Participants who reported any type of transportation-related access issue (compared to those without any difficulty traveling) had significantly higher satisfaction scores (β = 0.89; 95% CI: 0.06, 1.7).

**Figure 4.**
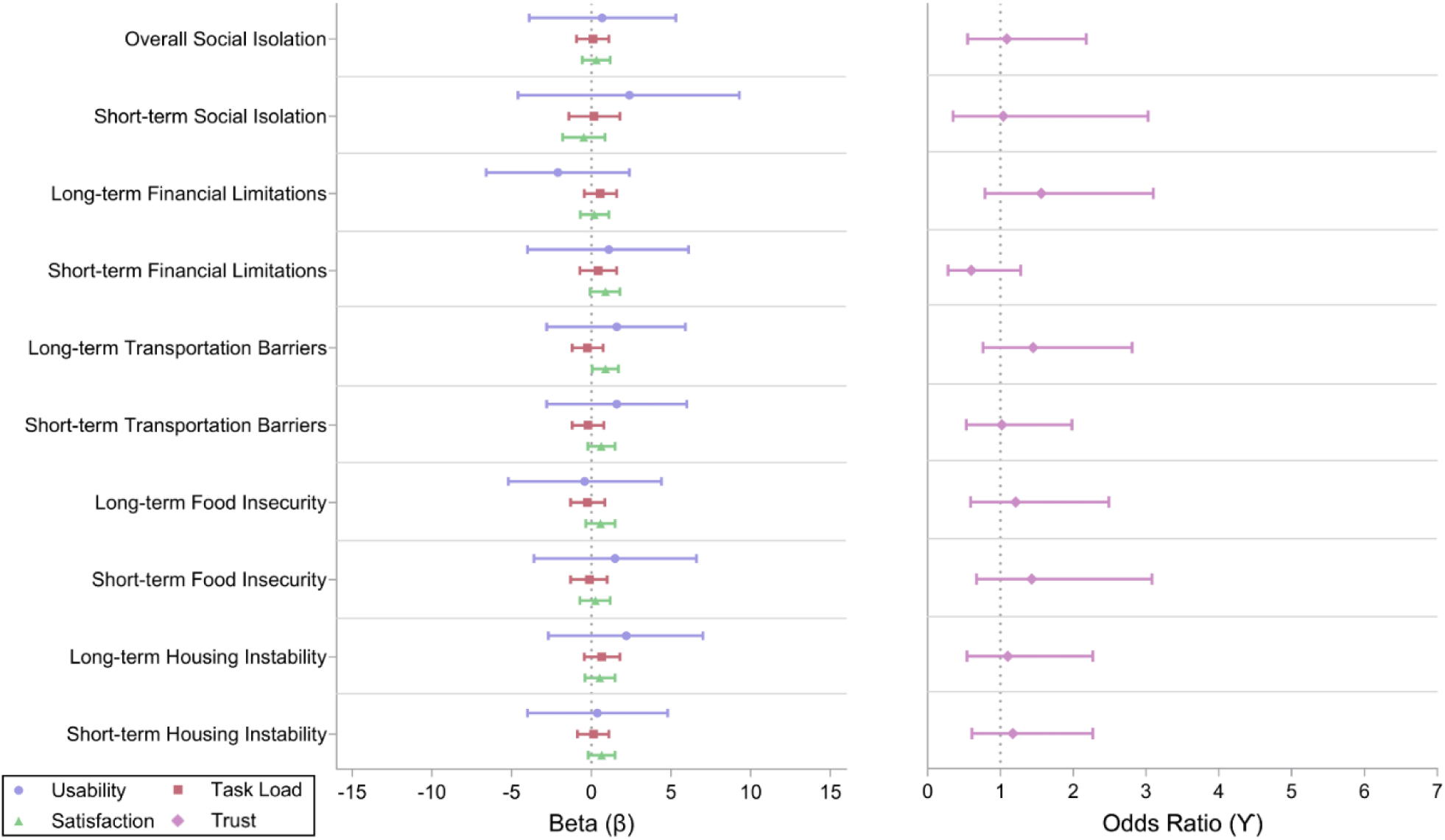
Regression analysis forest plot for HRSNs (binary) and user experience. First variable in each category is the reference value.

Several additional monotonic trends emerged that did not reach statistical significance but may warrant future exploration. The contrast for food insecurity (Often not enough to eat - Enough of the kinds of food we want to eat) showed a pronounced difference in estimated means for usability (Estimate = 9.59, SE = 5.18), and satisfaction (Estimate = 1.48, SE = 1.00), scaling down as food availability increased. Likewise, this contrast showed the most pronounced difference in task load (Estimate = −1.75, SE = 1.17), scaling up as food availability increased. Reports of utility shutoff followed a stepped pattern, with increasing levels of disruption (No → Yes → Already shut off) scaled positively with trust (Estimates for mean class scaling up from 3.00 to 3.33)). The contrast for affording utilities (Very Hard - Not hard at all) showed a pronounced difference in usability (Estimate = 1.72, SE = 3.34) and satisfaction (Estimate = 1.18, SE = 0.63), scaling down as difficulty decreased. Suggestive trends are presented in

### Supplementary Appendix

(Forest plots on Figures S8.5-S8.8; pairwise metrics on Tables S9.10-S9.20, and trust metrics on Tables S10.4-S10.7).

Meanwhile, in terms of notable non-monotonic shifts, a decrease in quantity of food eaten the household (RV: Enough food that we want to eat) trended with increased trust (Mean class estimate ranging from 2.95 to 3.30 compared to the mean class of 2.96 for ‘Enough of the kinds of food we want to eat’)). Reports of utility shutoff trend positively with task load (Estimated difference from 0.24 to 0.61 compared to no risk of shutoff)), and satisfaction (Estimated difference from 0 to 1.86 compared to no risk of shutoff). Lastly, increase in interactions with people they are close to trends positively with usability (Estimated difference from 0.13 to 4.11 compared to less than once a week) and trust (estimated mean class for 3.15 for 5+ times a week compared to 3.06 for less than once a week).

### Conversational analytics

To better understand user interactions and preferences, we analyzed chatbot conversation transcripts, examining message characteristics, interaction types, content specificity, and conversation topics. Since the interactions for scenarios 1-3 are scripted, our analysis in this section is only focused on conversational data during the free engagement (last scenario).

Participants generated a total of 204 chatbot transcripts through their engagements, where each user held between 1 to 4 unique conversations (M = 1.46, SD = 0.76). Participants interacted with the chatbot for approximately 3 minutes per session (M = 3 min 8 sec, SD = 4 min 59 sec), averaging 20 conversational turns (range: 3 to 49). Individual user messages averaged 3 words (17 characters), ranging from brief inputs (1 word, 3 characters) to longer statements (up to 43 words, 214 characters). Participants strongly preferred using assistive buttons (N = 2,749 button presses) rather than free-text entries (N = 1,151), with the ‘Coverage’ button most frequently used prior to session completion (see Table 1 in Supplementary Appendix 5).

Further qualitative examination of free-text entries provided deeper insights into how users communicated their resource needs. Using a structured rubric (Supplementary Appendix 3), two raters (AH and DIJ) assessed message specificity and user behaviors, achieving substantial agreement (Cohen’s Kappa; κ = 0.721). Users typically indicated specific community resources or facilities by name (M = 0.37, SD = 0.48), while a minority of the initiated interactions unrelated to social or resource needs (M = 0.91, SD = 0.28) (see Table 2 in Supplementary Appendix 5).

Keyword frequency analysis further illustrated user priorities (see **Table 3** in **Supplementary Appendix 5**), highlighting terms closely aligned with resource needs such as ‘program(s),’ (16%) ‘shelter,’ (12%) and ‘food’ (8%). Conversation topics similarly reflected key user concerns, predominantly focusing on financial issues (18%), housing (17%), and nutritional (16%) needs. Users also raised diverse unique topics beyond predefined HRSN categories, underscoring the chatbot’s capacity to address a broad range of community-related issues (see Table 4 in Supplementary Appendix 5). Including fallbacks in the conversation, chatbot successfully identified the user’s intent 1511 out of 1523 messages (99.21%).

### User Preferences

Participant free-text comments (open-ended responses to a positive experience, a negative experience, and suggested improvements for the chatbot) were analyzed and categorized into two components: the content and relevance of chatbot responses, and how information was presented and experienced during the interaction (see **Supplementary Appendix 6**). In their free-text feedback, participants discussed preferences for direct links, eligibility or scheduling information, and location-based customization. They also described the interface as intuitive and appreciated the chatbot’s speed. Furthermore, participants also desired voice-based interaction, more relevant local options, and improved mechanisms to refine their search early in the conversation. Sentiment analysis of the comments indicated generally positive feedback (M = 3.41, SD = 0.97), and intensity analysis showed moderate emotional expressiveness (M = 3.02, SD = 0.46). While many comments reflected satisfaction with ease of use and responsiveness, others noted limitations in personalization, specificity, and verbal communication capabilities.

After completion of the last scenario, nearly all participants (94%) reported the DAPHNE chatbot helpful in identifying relevant community resources. Post testing, they also reported preferred chatbot app enhancements (for future improvements beyond conversational capabilities). Participants expressed interest in several additional features, notably the inclusion of a “History and Favorites” (19%), the ability to create personal profiles (17%), password protection for enhanced security (15%), daily tips and personalized updates (15%), multilingual support (12%), integration with personal medical records (11%), and phone notifications (10%). A small proportion of users also suggested incorporating encouraging messaging, varied chatbot personalities, and additional technical guidance (see Supplementary Table 7).

## Discussion

The present mixed-methods evaluation sought to investigate user experience towards an AI-enabled chatbot (DAPHNE) that helped to identify unmet HRSNs and share real-time, community-based resource recommendations for the low income earning cohort. Building on our prior development,^18^ we assessed the usability, task load, satisfaction, and trust of DAPHNE with qualitative analyses of conversational transcripts and free-text feedback. In doing so, we offer one of the first empirical demonstrations that a rules-guided, LLM-supported chatbot can deliver just-in-time HRSN guidance with high acceptability (in terms of usability, satisfaction and trust) and minimal cognitive burden (task load).

### Demographic composition and reported HRSNs

The study sample was largely representative of the racially and ethnically diverse, low-income, caregiver population targeted by current U.S. screening recommendations. Nearly all participants reported at least one unmet HRSN, with food insecurity and financial strain predominating in the short term. The co-occurrence of multiple needs, including transportation barriers and chronic social isolation, underscores the layered vulnerabilities confronting these families and highlights the importance of integrated referral mechanisms rather than siloed interventions.^46,47^ Although online recruitment may select for digitally literate participants, the high prevalence of Medicaid coverage and bilingual status suggests that DAPHNE assessed by the cohort who are often underserved by traditional resource-navigation models.^48,49^

### User experiences

Across standardized instruments, the chatbot demonstrated high usability, low task load, satisfaction, and substantial perceived trustworthiness -metrics that surpass typical digital health benchmarks and are notable given the complexity of HRSN navigation.^49,50^ Conversational-analytics reinforce these quantitative ratings: users interacted through succinct, goal-oriented exchanges via text or button options, the dialogue manager mapped intents with high fidelity, and fallback LLM prompts were rarely invoked, all of which enabled transparent dialogue and align with the low task load observed.^51,52^

Qualitative feedback adds nuance, indicating that while participants appreciated the interface’s intuitiveness and speed, they sought richer informational specificity, such as detailed eligibility criteria, hyperlinked referrals, and finer geospatial granularity. Users also expressed interest in multimodal enhancements, including voice input, personalized dashboards, secure logins, and proactive notifications. Sentiment analysis confirmed a generally positive effect yet only moderate emotional engagement, suggesting room for more empathic and context-aware dialogue.^53–55^ Together, these behavioral and attitudinal data suggest not merely high perceived ease of use scores but an expectation for deeper personalization and multimodal interaction that can convert brief screening encounters into sustained, longitudinal support.^53^

### Relationship between Demographics, HRSN and User experience

Demographic and HRSN covariates revealed limited but informative associations with user experience outcomes. Usability and trust were broadly consistent across sex,^56^ age,^57^ and insurance status, suggesting wide acceptability across populations that are often underserved in digital health contexts.^58^ One of the clearest demographic patterns emerged in task load: non-married participants, including those who were divorced, never married, or unmarried, reported significantly lower task load compared to their married peers. One explanation is that unmarried individuals may be more accustomed to independently managing daily tasks and thus more familiar with digital tools, leading to lower perceived effort when using the chatbot. However, the relationship between marital status and technology-related task load remains underexplored and warrants further study.

Additionally, participants with lower income showed a directional trend toward reduced usability, however, participants with higher education levels also trended toward lower usability and higher task load. Negative trend between education level and usability contrasts with the current literature on perceptions towards digital health.^59,60^ One possible explanation is the overall low socioeconomic status of our sample, where limited variation in income and education may blunt the typical advantages associated with higher educational attainment. In this context, higher education may raise expectations for the chatbot’s capabilities, especially regarding access to resources, which, if unmet, could result in lower perceived usability. Similarly, participants already familiar with established ways of finding resources (digital or traditional) may perceive the chatbot as redundant or inefficient (e.g., extra steps of conversations), contributing to a higher task load. These findings suggest the chatbot may be particularly beneficial for individuals who are new to navigating resource systems. This interpretation is consistent with findings from Kocielnik et al.,^21^ who reported that emergency department patients with lower health literacy were more receptive to chatbot-based screening than those with higher literacy levels.

Several HRSNs were associated with user experience outcomes. Participants reporting transportation limitations or recent utility shutoffs (two direct indicators of material hardship) reported higher trust and satisfaction. This may reflect a heightened appreciation or perceived benefit for accessible digital tools when traditional health system access is disrupted, aligning with studies showing that resource-constrained users often assign greater value to mobile or remote interventions that reduce friction in service navigation.^61^ Users who reported being very often worried about food also reported lower task load, possibly suggesting that the chatbot was perceived as cognitively manageable during periods of stress, which is an important consideration given known links between food insecurity and reduced bandwidth for health engagement.^62^ Notably, individuals reporting limited social interaction (often used as a proxy for social isolation) showed a non-significant trend toward higher satisfaction, a finding that complicates assumptions in the literature that contributes to the growing literature of perceived relational or emotional utility of AI chatbots.^63–65^ These findings challenge the assumption that unmet needs uniformly impair digital engagement; instead, they suggest that digital tools may offer unique and sometimes amplified relevance for users navigating structural barriers.^66^

### Implications

Identification for HRSNs in clinical settings is a crucial first step, though often does not translate into successful resource connections for patients out of the clinic.^34,67^ Therefore, these results are encouraging. DAPHNE functioned not as a passive screener but as an active bridge to tailored referrals, directly addressing the long-standing “screen-to-referral” gap documented.^68,69^ From a design perspective, our findings imply that an effective social-care chatbot for low-earning cohorts should pursue quick, low-load button prompts while preserving free-text channels that capture nuanced circumstances. Equity-focused co-design involving community stakeholders and ongoing equity audits, tracking intent-recognition errors and unmet requests, will be critical to prevent digital access biases and safeguard equitable linkage to resources.^70,71^ Moreover, given that technology alone cannot resolve structural barriers, provider-technology collaboration, user education, and positioning chatbots as social-care navigators may further ensure culturally relevant and context-sensitive support, aligning closely with contemporary health-equity frameworks.^72^

Looking ahead, our study can initiate the discussions on the potential for AI-enabled chatbots to support policy mandates for HRSNs screening, identification and requirements of resource sharing. Given current CMS requirements for hospitals to screen and report social drivers of health and Medicaid initiatives incentivizing systematic HRSN assessment,^73^ AI-enabled chatbots could substantially ease implementation burdens.^74^ However, the USPSTF has so far found insufficient evidence to recommend universal screening for certain social risks (such as food insecurity) due to uncertain impact on health outcomes.^75^ This evidence gap highlights that simply implementing screening mandates is not enough, effective solutions are needed to actually address the identified needs. Our findings contribute to an emerging evidence base by illustrating that an AI-enabled chatbot, as a navigator, can achieve high engagement and potentially assist in resource connection. If such tools can be proven to improve outcomes (for example, reducing unmet needs or downstream health care utilization), they could substantially strengthen the case for routine social risk screening and intervention as a covered, reimbursable preventive service.^76^

### Strengths and Limitations

Key strengths of this study include the mixed-methods, convergent design and deployment of the chatbot in a realistic context with real-time resource data. Collecting quantitative (survey) and qualitative (interaction logs and interviews) data allowed us to triangulate findings, enhancing the validity of our conclusions.^77^ The use of an actual resource referral API (as opposed to a static or hypothetical list) increases the validity of our usability results and demonstrates technical feasibility for real-world integration. We also prioritized user-centered development with an equity lens, recruiting a diverse sample of low-income participants and incorporating their feedback into design refinements.

Nonetheless, several limitations warrant discussion. First, outcomes were measured via self-report instruments and short-term task performance in English language only; we did not directly measure long-term impacts on resource uptake or health outcomes including multiple languages. Participants’ subjective ratings of trust and satisfaction may have been influenced by social desirability or novelty effects. Second, the study lacked a comparator group, without a control condition (e.g. usual care or an alternative navigation method), we cannot definitively attribute benefits to the chatbot or assess its relative effectiveness. Future studies should consider randomized trials or quasi-experimental designs to compare chatbot-assisted social care navigation against standard practice. Third, our sample size and setting limit generalizability. All participants had at least some internet access and consented to engage with a digital tool; thus, our findings may not extend to those with severe digital literacy barriers or distrust of technology.^78,79^ This digital divide bias means additional strategies are needed to reach and support the most marginalized groups who might not engage with a chatbot.^80^ Chatbot was capable of providing guidance only based on single API based resource, and nuanced or complex HRSNs and guidance (such as, guidance to enroll support programs) may be lacking and affected perceived value. Finally, while our RAG-based approach was intended to ensure accuracy, we did not formally evaluate the accuracy of chatbot responses or recommendations. There remains a risk of hallucinations or errors when large language models are involved.^81^ Although no serious issues were reported in our study, establishing rigorous content validation and oversight (such as human review of a subset of chatbot outputs or user-friendly feedback mechanisms) will be important as the system scales.

### Conclusions

In conclusion, our study demonstrates the promise of an AI-enabled chatbot, DAPHNE, to efficiently identify HRSNs and connect families with community resources in an acceptable and user-friendly manner. It contributes to the growing literature on digital social care practices and integration by providing empirical evidence that a chatbot can engage vulnerable populations with high usability and trust. To fully realize this potential, future work must rigorously evaluate effectiveness, ensure equity in design and deployment, and address practical implementation challenges. With continued refinement and evidence generation, AI-driven chatbots could become a valuable adjunct in the ongoing efforts to integrate social care into healthcare.

## Supporting information

STROBE

## Data Availability

All data produced in the present study are available upon reasonable request to the authors

## Data Sharing

Due to the sensitive nature of participants’ conversations with the chatbot, we are unable to share individual-level transcripts and historical records of interactions.

## Ethics

This study received approval to conduct human-subjects research (STUDY00004369) from the Internal Review Board (FWA000002860) at Nationwide Children’s Hospital.

## Acknowledgements

We are thankful to Amazon Web Services for AWS Social Responsibility and Impact credits, and FindHelp.org for research fellowship towards granting cost-free community resource access via their API. We thank MEMOTEXT for their technical support on chatbot map UI elements. We acknowledge Ting Zhu for his constructive feedback about our research.

## Contributions

E.S. conceived the study, led its design, and supervised implementation, analysis, and manuscript development. E.S., D.I.J. and S.A.H. contributed equally to drafting and preparation of the manuscript. D.I.J. led data collection and, together with S.A.H., performed data cleaning, preparation, and analysis. D.I.J. also led data visualization and supported interpretation of results. C.R. led the statistical modeling and supported quantitative analysis. A.B.K., M.S., E.F.L., A.P.E., M.D., and A.P. provided domain expertise and contributed to the review and revision of the final manuscript. All authors reviewed and approved the final version of the paper.

