## Supplemental Appendix for "Evaluating User Experiences with an AI Chatbot for Health-Related Social Needs: A Cross-Sectional Mixed Methods Study"

### Supplementary Appendix 1

#### Scenario 1 of 4: Finding a food program

In this scenario 1, let's assume you need to find currently available nutrition support programs:

1. In response to the chatbot's question ("What are you looking for today?"), ask about a nutrition program (e.g., food pantries, soup kitchens, or home-delivered meals)
2. Type the zip code: 43201 into the chatbot and confirm.
3. Review suggested 3 resources and programs.
4. Read "More Details" of one of the programs suggested.
5. Check operating "Business Hours" of the program you selected.

#### Scenario 2 of 4: Giving feedback & learning cost

In this scenario 2, we will change the conversation towards finding health related resources:

1. Continue the previous conversation by clicking "I'm Done."
2. When asked if the latest recommendation helped, select "Yes". This feedback will be used to update our resources and improve our responses for our users the next time.
3. Then, the chatbot asks if any other help is needed, click or type "Yes." to ask for other resources.
4. Select or type for "health-related" resources and then enter new zip code: 43215
5. Expand the suggestions by clicking on "More Resources"
6. Pick one of the resources or programs suggested by clicking "More Details" and read the information.
7. Ask about the program's "Cost" and read the information.
8. Click on "Go Back" to list again the resources.

#### Scenario 3 of 4: Housing

In this scenario 3, we will explore housing-related supporting programs:

1. Continue the previous conversation and click on "Change My Location"
2. Enter zip code: 45402 and review the suggested resources.
3. Click on "Find Help for Something Else" and type to ask about shelter and residential programs.

4. Click on "Contact Info" of any suggested program and review contact information, and then ask about their "Coverage."

#### Scenario 4 of 4: Exploration

In this scenario 4, you will be able to use the chatbot as you prefer:

1. Click on "I'm Done", then follow conversation to agree or disagree that you found what you were looking for, and then click "Yes" to the question of "Is there anything else I can help with?"
2. In this new session, explore the chatbot freely. Search for any community resources for yourself or someone you know with any zip code.
    - You may compare programs with other nearby zip codes.
    - You may try viewing the location of a resource using the "Open Maps" button.

### Supplementary Appendix 2

#### Rubric for Sentiment and Intensity

**Sentiment Category (1–5)**

- **1 (Negative)**: Strong dissatisfaction, frustration, confusion, disappointment, or harsh criticism
- **2 (Somewhat Negative)**: Mild dissatisfaction, frustration, or confusion
- **3 (Neutral)**: Factual statements without distinct emotion; balanced or mixed with little to no affect
- **4 (Somewhat Positive)**: Mild satisfaction or generally positive feelings
- **5 (Positive)**: Strong satisfaction, appreciation, happiness, or clear success

**Intensity Level (1–5)**

- **1 (Low)**: Minimal or negligible emotional content
- **2 (Moderately Low)**: Mild emotional content, somewhat subtle expressions of sentiment
- **3 (Medium)**: Moderate emotional expression, neither extreme nor minimal
- **4 (Moderately High)**: Noticeable emotional language but not overwhelmingly strong
- **5 (High)**: Very strong emotional language, clearly intense (e.g., “extremely,” “fantastic,” “terrible”)

**Application Guidelines**

1. **Provide Two Ratings:**

   For each user response, give both a "Sentiment Category" rating (1–5) and an "Intensity Level" rating (1–5), along with a brief explanation.

2. **Handling Borderline Cases:**

   If it's unclear whether to classify feedback as 1 (Negative) or 2 (Somewhat Negative), or 4 (Somewhat Positive) or 5 (Positive), lean toward 2 or 4 unless the feedback clearly belongs in the most negative or most positive category.

   Use 3 (Neutral) for factual, balanced statements or moderate emotional expression.

3. **Mixed/Conflicting Sentiment:**

   If a response shows both positive and negative aspects with different intensities, select the **more extreme** sentiment category (e.g., if a user mostly praises but includes one strongly critical point, the negative might dominate if it's clearly intense).

### Supplementary Appendix 3

#### Rubric for Specificity and Behaviors

Score each category as either:

- 1 (Present): The feature is clearly included
- 0 (Absent): The feature is missing or too vague

Use the user's message and chatbot prompt as context for evaluation.

##### (1) Mentions of a Specific Resource

Score 1 if the participant names something more precise than a general category

Examples:

- Specific organizations or programs (e.g., "Medicaid", "Salvation Army")
- Subsets or qualifiers (e.g., "allergen-free food", "bus tokens", "computer classes")

Score 0 if the response is generic or vague

Examples:

- "food help"
- "housing support"

Synonyms that restate a broad domain without narrowing it

Generic domain list: food, housing, goods, transit, health, money, care, education, work, legal

##### (2) Mentions of the Purpose for the Resource

Score 1 if the user explains who the help is for or why they need it

Examples:

- “for my fiancée”
- “so I can get to work”
- “for my friend who’s diabetic”

Score 0 if there’s no purpose or only a vague plea

Example: “I just need help”

##### (3) Mentions their Personal Background

Score 1 if the user shares any detail about their life situation

Examples:

- “I just moved”
- “I have 3 kids”
- “I lost my job”

Score 0 if there's no personal context provided.

##### (4) Mentions Multiple Needs

Score 1 if the user identifies two or more distinct needs that require different types of assistance

Examples:

- From different domains (e.g., food and housing), or
- From the same domain but covering different timescales or scopes (e.g., groceries today vs. long-term SNAP)

Score 0 if only one need is mentioned, or multiple phrases point to the same issue

Examples:

- “place to stay” + “safe housing”
- “rent help” + “finding an apartment”

##### (5) Mentions Thoughts on Finding a Resource

Score 1 if the user shows any interest in getting help or locating a resource

Examples:

- "I'm overwhelmed and don't know what to do"
- "Is there a place I can go?"

Score 0 if the message is unrelated to help-seeking

Examples:

- Jokes
- Meta-comments ("This app is weird")
- Disengagement ("idk")

### Supplementary Appendix 4

#### Table S4.1. Self-reported health-related social needs

| **Health-related Social Needs** | **N** | **%** |
|---|---|---|
| *Within the past 12 months, have you ever stayed: outside, in a car, in a tent, in an overnight shelter, or temporarily in someone else's home (i.e. couch-surfing)?* | | |
| Yes* | 36 | 28% |
| No | 92 | 72% |
| *In the last month, have you had concerns about the condition or quality of your housing?* | | |
| Yes* | 59 | 46% |
| No | 69 | 54% |
| *Within the past 12 months, have you worried that your food would run out before you got money to buy more?* | | |
| Never | 37 | 29% |
| Sometimes* | 58 | 45% |
| Often* | 19 | 15% |
| Very Often* | 14 | 11% |
| *Which of these statements best describes the food eaten in your household in the last month?* | | |
| Often not enough to eat* | 7 | 5% |
| Sometimes not enough to eat* | 17 | 13% |
| Enough but not always the kinds of food we want* | 73 | 57% |
| Enough of the kinds of food we want to eat | 31 | 25% |
| *In the past 12 months, has lack of transportation kept you from medical appointments, meetings, work, or from getting things needed for daily living? *** | | |
| Yes, it has kept me from medical appointments or getting medications* | 34 | 23% |
| Yes, it has kept me from non-medical meetings, appointments, work, or getting things that I needed* | 48 | 32% |
| No | 66 | 45% |

| | | |
|---|---|---|
| *In the last month, did you put off or neglect going somewhere because of distance or transportation?* | | |
| Yes* | 73 | 57% |
| No | 55 | 43% |
| *In the past 12 months, has the electric, gas, oil, or water company threatened to shut off services in your home?* | | |
| No | 79 | 62% |
| Yes* | 44 | 34% |
| Already Shut Off* | 5 | 4% |
| *In the last month, how hard was it for you to pay for the very basics like food, housing, medical care, and heating?* | | |
| Not hard at all | 32 | 25% |
| Somewhat Hard* | 71 | 55% |
| Very hard* | 25 | 20% |
| *How often do you need to have someone help you when you read instructions, pamphlets, or other written material?* | | |
| Never | 102 | 80% |
| Rarely | 13 | 10% |
| Sometimes* | 11 | 8% |
| Often* | 1 | 1% |
| Always* | 1 | 1% |
| *How confident are you filling out medical forms by yourself?* | | |
| Extremely | 82 | 64% |
| Very much | 34 | 27% |
| Slightly* | 9 | 7% |
| Slightly not* | 0 | 0% |
| Very little* | 2 | 2% |
| Not at all* | 1 | 1% |
| *How often do you feel isolated from others?* | | |
| Hardly ever | 45 | 35% |
| Some of the time* | 49 | 38% |
| Often* | 34 | 27% |
| *How often do you see or talk to people that you care about and feel close to? (For example: Talk to friends over the phone, visiting friends or family, going to church or club meetings)* | | |
| 5 or more time a week | 44 | 34% |
| 3 to 5 times a week | 34 | 27% |
| 1 or 2 times a week | 36 | 28% |
| Less than once a week* | 14 | 11% |

** Marked answers are positive screenings of HRSNs for binary scoring*
*** Not mutually exclusive, as multiple transportation needs were captured here*

#### Table S4.2. Statistical regressions between demographics and user experience

| | Usability | | Trust | | Task load | | Satisfaction | |
|---|---|---|---|---|---|---|---|---|
| **Characteristic** | **Beta** | **95% CI** | **OR** | **95% CI** | **Beta** | **95% CI** | **Beta** | **95% CI** |
| **Sex** | | | | | | | | |
| Female | 0.00 | — | 1.00 | — | 0.00 | — | 0.00 | — |
| Male | -0.20 | -4.9, 4.5 | 1.28 | 0.64, 2.57 | 0.65 | -0.40, 1.7 | 0.31 | -0.59, 1.2 |
| **Age** | | | | | | | | |
| Young Adults | 0.00 | — | 1.00 | — | 0.00 | — | 0.00 | — |
| Middle-Aged Adults | 0.25 | -4.3, 4.7 | 0.82 | 0.42, 1.60 | -0.64 | -1.7, 0.39 | 0.99 | 0.14, 1.8 |
| Senior Adults | -14 | -31, 3.8 | 2.47 | 0.18, 59.5 | 0.15 | -3.9, 4.2 | -2.7 | -6.0, 0.63 |
| **Education** | | | | | | | | |
| Highschool | 0.00 | — | 1.00 | — | 0.00 | — | 0.00 | — |
| *Higher Education** | *-2.2* | *-7.6, 3.1* | *1.3* | *0.58, 2.93* | *-0.21* | *-1.4, 1.0* | *0.15* | *-0.88, 1.2* |
| Some college, no degree | 1.0 | -5.0, 7.1 | 1.89 | 0.75, 4.82 | -1.3 | -2.6, 0.06 | 0.67 | -0.51, 1.9 |
| Associate degree | -3.7 | -11, 3.3 | 1.06 | 0.37, 3.04 | -0.32 | -1.9, 1.2 | -0.30 | -1.7, 1.1 |
| Bachelor’s degree | -5.9 | -13, 0.79 | 0.90 | 0.32, 2.54 | 0.89 | -0.58, 2.4 | -0.31 | -1.6, 1.0 |
| Master’s degree | -4.9 | -14, 4.5 | 1.19 | 0.27, 5.21 | 1.9 | -0.16, 4.0 | -0.11 | -1.9, 1.7 |
| Other | 3.6 | -11, 18 | 0.98 | 0.10, 11.1 | -0.83 | -4.1, 2.4 | 1.1 | -1.8, 4.0 |
| **Income** | | | | | | | | |
| Less than $10,000 | 0.00 | — | 1.00 | — | 0.00 | — | 0.00 | — |
| *$10,000-$39,000** | *-2.2* | *-7.6, 3.1* | *1.3* | *0.58, 2.93* | *-0.21* | *-1.4, 1.0* | *0.15* | *-0.88, 1.2* |
| $10,000-$19,999 | -5.0 | -13, 3.5 | 0.80 | 0.22, 2.91 | 0.52 | -1.4, 2.5 | -0.79 | -2.4, 0.85 |
| $20,000-$29,999 | -2.2 | -9.1, 4.6 | 2.03 | 0.71, 5.90 | 0.97 | -0.60, 2.5 | -0.30 | -1.6, 1.0 |
| $30,000-$39,999 | 0.40 | -6.2, 7.0 | 1.74 | 0.64, 4.74 | 0.92 | -0.57, 2.4 | 0.34 | -0.93, 1.6 |
| **Do you speak a language other than English at home?** | | | | | | | | |
| No | 0.00 | — | 1.00 | — | 0.00 | — | 0.00 | — |
| Yes | -2.7 | -7.9, 2.6 | 0.82 | 0.38, 1.77 | 0.44 | -0.75, 1.6 | 0.22 | -0.80, 1.2 |
| **Currently employed** | | | | | | | | |

| | Usability | | Trust | | Task load | | Satisfaction | |
|---|---|---|---|---|---|---|---|---|
| **Characteristic** | **Beta** | **95% CI** | **OR** | **95% CI** | **Beta** | **95% CI** | **Beta** | **95% CI** |
| No | 0.00 | — | 1.00 | — | 0.00 | — | 0.00 | — |
| Yes | 1.8 | -2.6, 6.3 | 1.50 | 0.77, 2.93 | 0.38 | -0.63, 1.4 | 0.80 | -0.05, 1.6 |
| **Insurance** | | | | | | | | |
| No | 0.00 | — | 1.00 | — | 0.00 | — | 0.00 | — |
| Yes | 0.77 | -5.8, 7.4 | 2.62 | 1.03, 6.76 | -0.46 | -2.0, 1.0 | 0.51 | -0.76, 1.8 |
| **Marital Status** | | | | | | | | |
| Married | 0.00 | — | 1.00 | — | 0.00 | — | 0.00 | — |
| *Other Partnerships** | *2.8* | *-1.7, 7.3* | *0.89* | *0.45, 1.74* | *-1.5* | *-2.5, -0.46* | *0.01* | *-0.86, 0.89* |
| Divorced | 5.7 | -2.8, 14 | 0.51 | 0.15, 1.78 | -2.7 | -4.6, -0.85 | 0.60 | -1.1, 2.3 |
| Widowed | -17 | -42, 7.9 | 0.09 | 0.00, 2.86 | 0.63 | -4.9, 6.1 | -3.0 | -7.8, 1.8 |
| Separated | 2.6 | -10, 15 | 0.72 | 0.11, 5.04 | -0.78 | -3.6, 2.1 | 0.25 | -2.2, 2.7 |
| Never Married | 1.7 | -3.5, 6.8 | 0.81 | 0.37, 1.76 | -1.3 | -2.4, -0.14 | -0.20 | -1.2, 0.80 |
| Unmarried Couple | 4.7 | -1.6, 11 | 1.65 | 0.63, 4.43 | -1.4 | -2.8, 0.00 | 0.27 | -0.96, 1.5 |
| **Current number of children under your care** | | | | | | | | |
| None | 0.00 | — | 1.00 | — | 0.00 | — | 0.00 | — |
| *Any Child Dependents** | *6.9* | *-5.6, 19* | *4.01* | *0.70, 22.9* | *-1.4* | *-4.3, 1.4* | *0.27* | *-2.2, 2.7* |
| 1-2 | 6.6 | -6.0, 19 | 3.86 | 0.67, 22.2 | -1.4 | -4.2, 1.5 | 0.14 | -2.3, 2.6 |
| 3-4 | 7.5 | -5.9, 21 | 4.53 | 0.68, 30.1 | -1.7 | -4.8, 1.4 | 0.66 | -1.9, 3.2 |
| 5 or more | 16 | -5.3, 37 | 10.2 | 0.47, 33.9 | -1.3 | -6.2, 3.5 | 2.3 | -1.9, 6.4 |
| **Do you or your dependent child have special healthcare needs or chronic conditions?** | | | | | | | | |
| No | 0.00 | — | 1.00 | — | 0.00 | — | 0.00 | — |
| Yes | 2.9 | -1.7, 7.5 | 1.48 | 0.74, 3.01 | -0.62 | -1.7, 0.43 | 0.64 | -0.26, 1.5 |
| **Prior use of chatbots or virtual assistants** | | | | | | | | |
| No | 0.00 | — | 1.00 | — | 0.00 | — | 0.00 | — |
| Yes | -3.8 | -13, 5.7 | 0.68 | 0.17, 2.60 | 0.32 | -1.9, 2.5 | -0.29 | -2.1, 1.6 |

| | Usability | | Trust | | Task load | | Satisfaction | |
|---|---|---|---|---|---|---|---|---|
| **Characteristic** | **Beta** | **95% CI** | **OR** | **95% CI** | **Beta** | **95% CI** | **Beta** | **95% CI** |
| **Frequency of chatbot or virtual assistant use** | | | | | | | | |
| Infrequent (<2 times per week) | 0.00 | — | 1.00 | — | 0.00 | — | 0.00 | — |
| *Frequent (≥2 times per week)** | *2.4* | *-2.4, 7.3* | *1.66* | *0.81, 3.42* | *0.28* | *-0.83, 1.4* | *0.45* | *-0.47, 1.4* |
| Occasional (2-6 times per week) | 3.5 | -2.0, 9.0 | 1.93 | 0.85, 4.43 | -0.29 | -1.5, 0.95 | 0.66 | -0.39, 1.7 |
| Often (>7 times per week) | 1.3 | -4.4, 6.9 | 1.41 | 0.61, 3.25 | 0.94 | -0.34, 2.2 | 0.21 | -0.86, 1.3 |

Abbreviations: CI = Confidence Interval, OR = Odds Ratio

* = Binarized comparison of means and confidence intervals between the reference point and all other possible characteristics per measure

#### Table S4.3. Statistical regressions between health-related social needs and user experiences

| | Usability | | Trust | | Task load | | Satisfaction | |
|---|---|---|---|---|---|---|---|---|
| **Characteristic** | **Beta** | **95% CI** | **OR** | **95% CI** | **Beta** | **95% CI** | **Beta** | **95% CI** |
| **Within the last 12 months, have you ever stayed outside, in a car/tent/overnight shelter, or temporarily in someone else's home?** | | | | | | | | |
| No | 0.00 | — | 1.00 | — | 0.00 | — | 0.00 | — |
| Yes | 2.2 | -2.7, 7.0 | 1.10 | 0.54, 2.27 | 0.66 | -0.44, 1.8 | 0.53 | -0.40, 1.5 |
| **In the last month, any concerns about quality of housing?** | | | | | | | | |
| No | 0.00 | — | 1.00 | — | 0.00 | — | 0.00 | — |
| Yes | 0.39 | -4.0, 4.8 | 1.17 | 0.61, 2.27 | 0.14 | -0.86, 1.1 | 0.65 | -0.19, 1.5 |
| **Within the last 12 months, have you worried that your food would run out before you got money to buy more?** | | | | | | | | |
| No | 0.00 | — | 1.00 | — | 0.00 | — | 0.00 | — |
| *Yes** | *-0.42* | *-5.2, 4.4* | *1.21* | *0.59, 2.49* | *-0.25* | *-1.3, 0.85* | *0.58* | *-0.34, 1.5* |
| Sometimes | -0.63 | -5.9, 4.6 | 1.36 | 0.62, 2.96 | -0.07 | -1.2, 1.1 | 0.46 | -0.54, 1.5 |
| Often | -1.3 | -8.3, 5.7 | 0.75 | 0.26, 2.15 | 0.40 | -1.2, 2.0 | 0.30 | -1.0, 1.6 |

| | Usability | | Trust | | Task load | | Satisfaction | |
|---|---|---|---|---|---|---|---|---|
| **Characteristic** | **Beta** | **95% CI** | **OR** | **95% CI** | **Beta** | **95% CI** | **Beta** | **95% CI** |
| Very Often | 1.6 | -6.2, 9.4 | 1.44 | 0.48, 4.43 | -1.8 | -3.6, -0.11 | 1.5 | -0.01, 3.0 |
| **Quantity of food eaten in your household within the last month:** | | | | | | | | |
| Enough of the kinds of food we want to eat | 0.00 | — | 1.00 | — | 0.00 | — | 0.00 | — |
| *Not enough or kinds of food** | *1.5* | *-3.6, 6.6* | *1.43* | *0.67, 3.08* | *-0.11* | *-1.3, 1.0* | *0.27* | *-0.72, 1.2* |
| Enough but not always the kinds of food we want | 0.59 | -4.7, 5.8 | 1.36 | 0.61, 3.01 | 0.22 | -0.97, 1.4 | 0.06 | -0.96, 1.1 |
| Sometimes not enough to eat | 2.0 | -5.4, 9.4 | 2.32 | 0.75, 7.42 | -0.87 | -2.5, 0.81 | 0.66 | -0.77, 2.1 |
| Often not enough to eat | 9.6 | -0.66, 20 | 0.98 | 0.25, 3.93 | -1.7 | -4.1, 0.58 | 1.5 | -0.50, 3.5 |
| **Has lack of transportation impacted your ability to get places?** | | | | | | | | |
| No | 0.00 | — | 1.00 | — | 0.00 | — | 0.00 | — |
| *Yes** | *1.6* | *-2.8, 5.9* | *1.45* | *0.76, 2.81* | *-0.25* | *-1.2, 0.74* | *0.89* | *0.06, 1.7* |
| Yes, both medical and non-medical appointments | -0.01 | -6.3, 6.3 | 1.04 | 0.42, 2.63 | -0.59 | -2.0, 0.85 | 1.1 | -0.08, 2.3 |
| Yes, medical appointments/medications | 1.4 | -5.9, 8.7 | 3.21 | 1.03, 10.7 | -0.20 | -1.9, 1.5 | 1.1 | -0.32, 2.5 |
| Yes, non-medical meetings/appointments | 2.8 | -2.8, 8.4 | 1.30 | 0.56, 3.05 | -0.04 | -1.3, 1.2 | 0.64 | -0.42, 1.7 |
| **In the last month, did you put off or neglect going somewhere due to distance or lack of transportation?** | | | | | | | | |
| No | 0.00 | — | 1.00 | — | 0.00 | — | 0.00 | — |
| Yes | 1.6 | -2.8, 6.0 | 1.02 | 0.53, 1.98 | -0.21 | -1.2, 0.79 | 0.62 | -0.22, 1.5 |
| **In the past 12 months, has the electric, gas, oil, or water company threatened to shut off services in your home?** | | | | | | | | |
| No | 0.00 | — | 1.00 | — | 0.00 | — | 0.00 | — |
| *Yes, or already shut off** | *-2.1* | *-6.6, 2.4* | *1.56* | *0.79, 3.10* | *0.57* | *-0.45, 1.6* | *0.19* | *-0.68, 1.1* |
| Yes | -3.1 | -7.7, 1.5 | 1.48 | 0.73, 3.03 | 0.61 | -0.45, 1.7 | 0.00 | -0.89, 0.88 |
| Already shut off | 6.9 | -4.3, 18 | 2.30 | 0.45, 13.0 | 0.24 | -2.3, 2.8 | 1.9 | -0.32, 4.0 |
| **In the last month, how hard was it for you to pay for the very basics like food, housing, medical care, and heating?** | | | | | | | | |

| | Usability | | Trust | | Task load | | Satisfaction | |
|---|---|---|---|---|---|---|---|---|
| **Characteristic** | **Beta** | **95% CI** | **OR** | **95% CI** | **Beta** | **95% CI** | **Beta** | **95% CI** |
| Not hard | 0.00 | — | 1.00 | — | 0.00 | — | 0.00 | — |
| *Hard** | *1.1* | *-4.0, 6.1* | *0.6* | *0.28, 1.28* | *0.43* | *-0.71, 1.6* | *0.89* | *-0.07, 1.8* |
| Somewhat hard | 0.83 | -4.4, 6.1 | 0.55 | 0.24, 1.21 | 0.81 | -0.36, 2.0 | 0.78 | -0.22, 1.8 |
| Very hard | 1.7 | -4.9, 8.3 | 0.76 | 0.28, 2.05 | -0.63 | -2.1, 0.84 | 1.2 | -0.08, 2.4 |
| **How often do you feel isolated from others?** | | | | | | | | |
| No (Hardly ever) | 0.00 | — | 1.00 | — | 0.00 | — | 0.00 | — |
| *Yes** | *0.69* | *-3.9, 5.3* | *1.09* | *0.55, 2.18* | *0.11* | *-0.93, 1.1* | *0.32* | *-0.56, 1.2* |
| Some of the time | -0.75 | -5.8, 4.3 | 1.26 | 0.58, 2.73 | 0.16 | -1.0, 1.3 | 0.12 | -0.86, 1.1 |
| Often | 2.8 | -2.8, 8.4 | 0.89 | 0.38, 2.06 | 0.04 | -1.2, 1.3 | 0.61 | -0.47, 1.7 |
| **How often do you see or talk to people that you care about and feel close to?** | | | | | | | | |
| 5 or more times a week | 0.00 | — | 1.00 | — | 0.00 | — | 0.00 | — |
| 3 to 5 times a week | -4.0 | -9.6, 1.7 | 0.74 | 0.32, 1.71 | 0.31 | -0.97, 1.6 | -0.43 | -1.5, 0.66 |
| 1 or 2 times a week | -1.8 | -7.3, 3.7 | 0.74 | 0.32, 1.67 | -0.56 | -1.8, 0.70 | -0.16 | -1.2, 0.91 |
| *At least once a week** | *2.4* | *-4.6, 9.3* | *1.04* | *0.35, 3.03* | *0.17* | *-1.4, 1.8* | *-0.47* | *-1.8, 0.87* |
| Less than once a week | -4.1 | -12, 3.5 | 0.80 | 0.25, 2.54 | -0.25 | -2.0, 1.5 | 0.29 | -1.2, 1.8 |

Abbreviations: CI = Confidence Interval, OR = Odds Ratio

* = Binarized comparison of means and confidence intervals between the reference point and all other possible characteristics per measure

### Supplementary Appendix 5

#### Table S5.1. Frequency of assistive button before leaving a chat session

| Assistive Button | N |
|---|---|
| Coverage | 34 |
| Find Help for Something Else | 31 |
| More Resources | 30 |
| Cost | 26 |
| Business Hours | 20 |

| | |
|---|---|
| Availability | 16 |
| Change My Location | 2 |

#### Table S5.2. Patterns in resource-seeking behaviors with specificity

| Category | Specific Resource | Purpose for the Resource | Personal Background | Multiple Needs | Thoughts on Finding a Resource |
|---|---|---|---|---|---|
| Mean (SD) | 0.37 (0.48) | 0.02 (0.13) | 0.03 (0.16) | 0.07 (0.26) | 0.91 (0.28) |
| Median | 0 | 0 | 0 | 0 | 1 |
| IQR | 1 | 0 | 0 | 0 | 0 |

#### Table S5.3. Frequencies of keywords in free text messages

| Word | N |
|---|---|
| program(s) | 217 |
| shelter(s) | 118 |
| residential | 108 |
| food | 80 |
| nutrition(al) | 76 |
| looking | 47 |
| related | 38 |
| pantry(ies) | 37 |
| health | 35 |
| resource(s) | 35 |

#### Table S5.4. Topics of interest

| Topic | N |
|---|---|
| money | 31 |
| housing | 30 |
| food | 28 |
| work | 14 |
| transit | 10 |
| education | 9 |
| goods | 7 |
| health | 6 |
| legal | 3 |

| | |
|---|---|
| care | 2 |
| *(other unique topics)* | *30* |

### Supplementary Appendix 6

#### Table S6.1. Qualitative domains and themes

| Domain | Theme | Quote | Participant |
|---|---|---|---|
| Knowledge | Use of links & follow-up | "I can use the phone numbers or links to follow up later on this resource." | 24 |
| | Scheduling & eligibility | "I want to make sure the resource I choose is open when I can get there." | 95 |
| | Location preferences | "If it's too far, then I want to adjust the range like other apps do." | 106 |
| Delivery | Interface usability | "The interface and language were very intuitive and simple." | 45 |
| | Response speed | "It was speedy and quick in response to my many questions." | 37 |
| | Voice interaction | "Maybe it could break it down for me by communicating via voice." | 18 |
| | Resource fit evaluation | "It kinda sorted out my needs for me." | 91 |
| Positive Feedback | Ease of use | "The chatbot was easy to use." | 73 |
| | Resource richness | "I loved how I was provided with more than 2 services." | 115 |
| | Guidance | "I like how it gave suggestions on info to look up." | 9 |
| Negative Feedback | Relevance issues | "Some of the resources listed were not very helpful for me and other family members." | 126 |
| | Voice absence | "The only negative I have is that the chatbot couldn't talk to me aloud." | 56 |
| | Location mismatch | "The resource results included places that wasn't in my area of interest." | 22 |
| Suggestions | Clarify search process | "It should help me reduce my queries to a question rather than many." | 90 |
| | Age-targeted suggestions | "It could provide options that are better suited for young adults." | 67 |
| | Refine prompts | "It should start by helping me be more specific when typing a question." | 3 |

### Supplementary Appendix 7

#### Table S7.1. Chatbot helpfulness and feature requests

| **Resource Helpfulness** | **N** | **%** |
|---|---|---|
| Yes, helpful for me or someone else. | 120 | 94% |
| No, unhelpful for me or someone else. | 8 | 6% |
| **Desired App Features** | | |
| History & Favorites Tab | 87 | 19% |
| Profile Creation | 80 | 17% |
| Password Protected App | 70 | 15% |
| Daily Tips & Personalized Updates | 69 | 15% |
| Multilingual Support | 57 | 12% |
| Connections to Your Medical Records | 50 | 11% |
| Phone Notifications | 48 | 10% |
| Other Features | 4 | 1% |
| Words of Encouragement | 1 | |
| Chatbot Personalities | 1 | |
| Technical Guidance | 2 | |

### Supplementary Appendix 8

#### Figure S8.1 Demographics and Usability Forest Plot

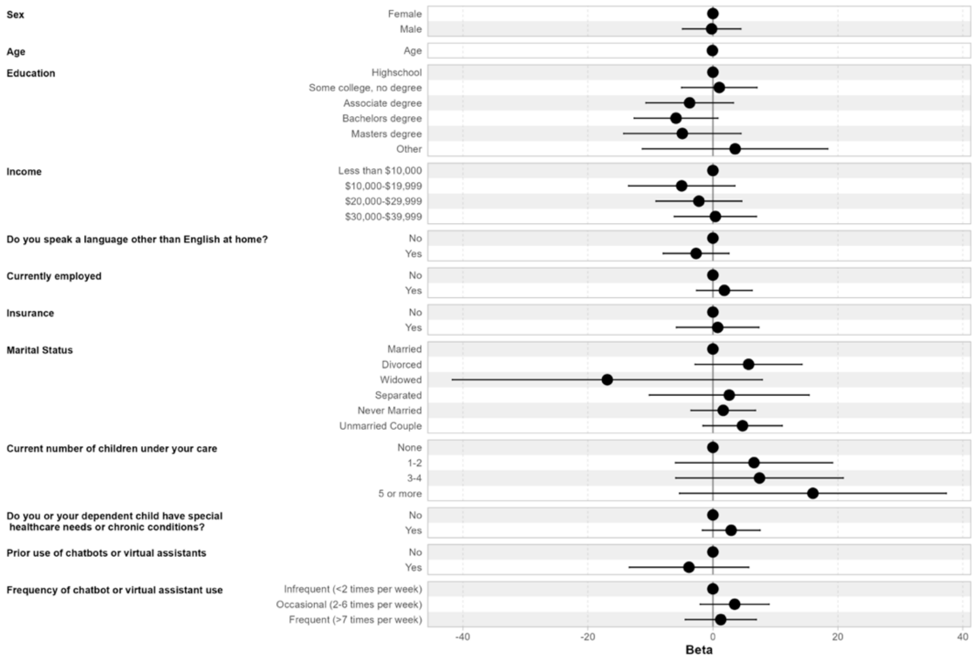


#### Figure S8.2 Demographics and Trust Forest Plot

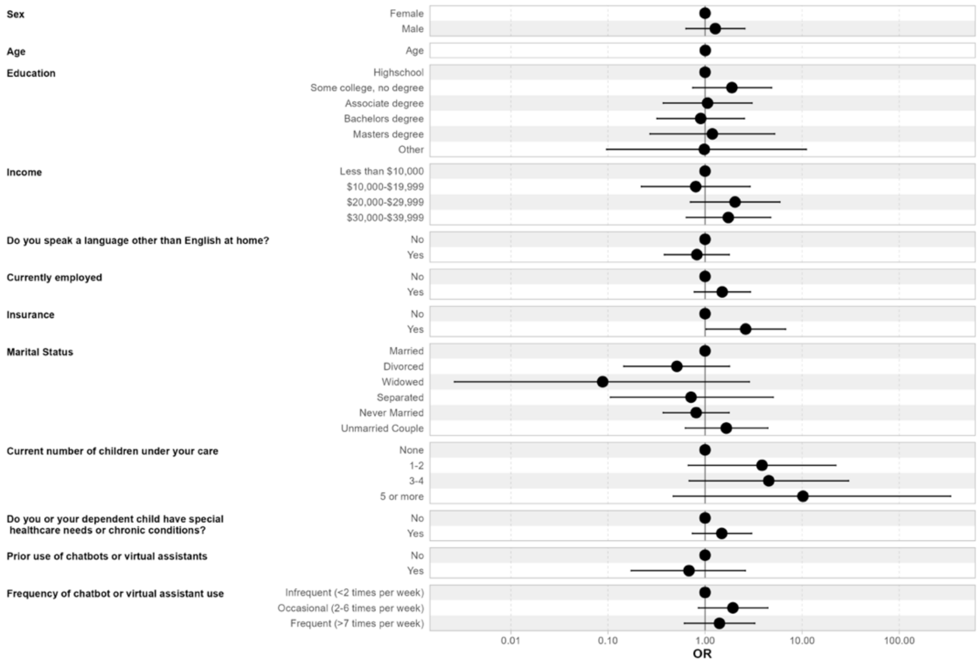

### Figure S8.3 Demographics and Task Load Forest Plot

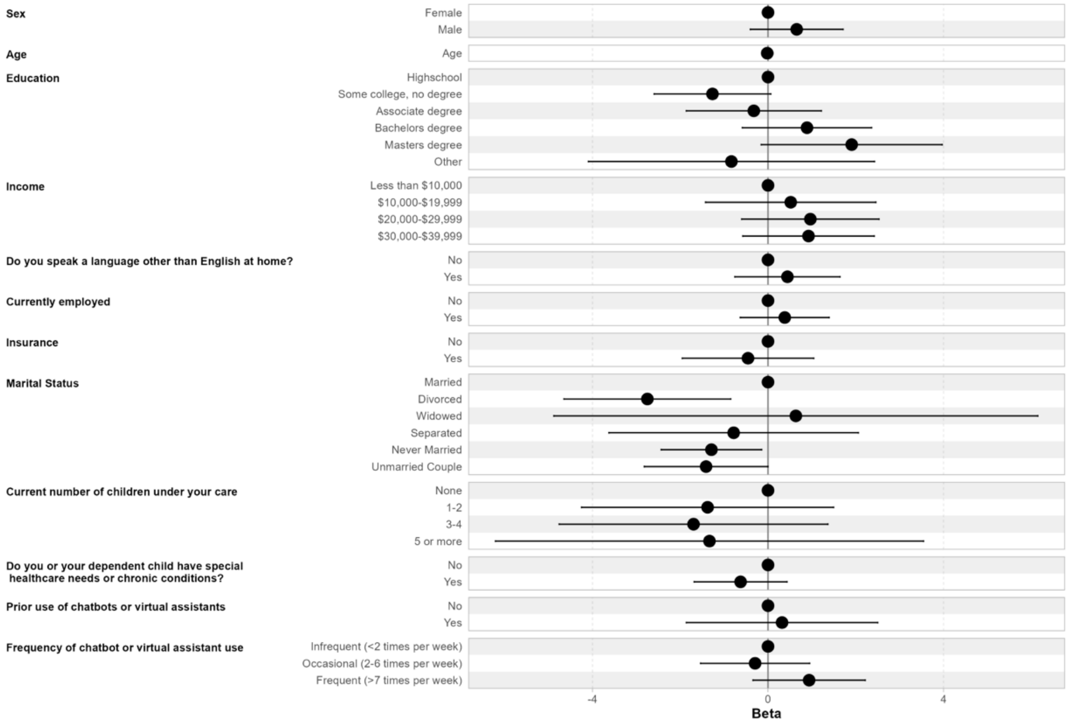


### Figure S8.4 Demographics and Satisfaction Forest Plot

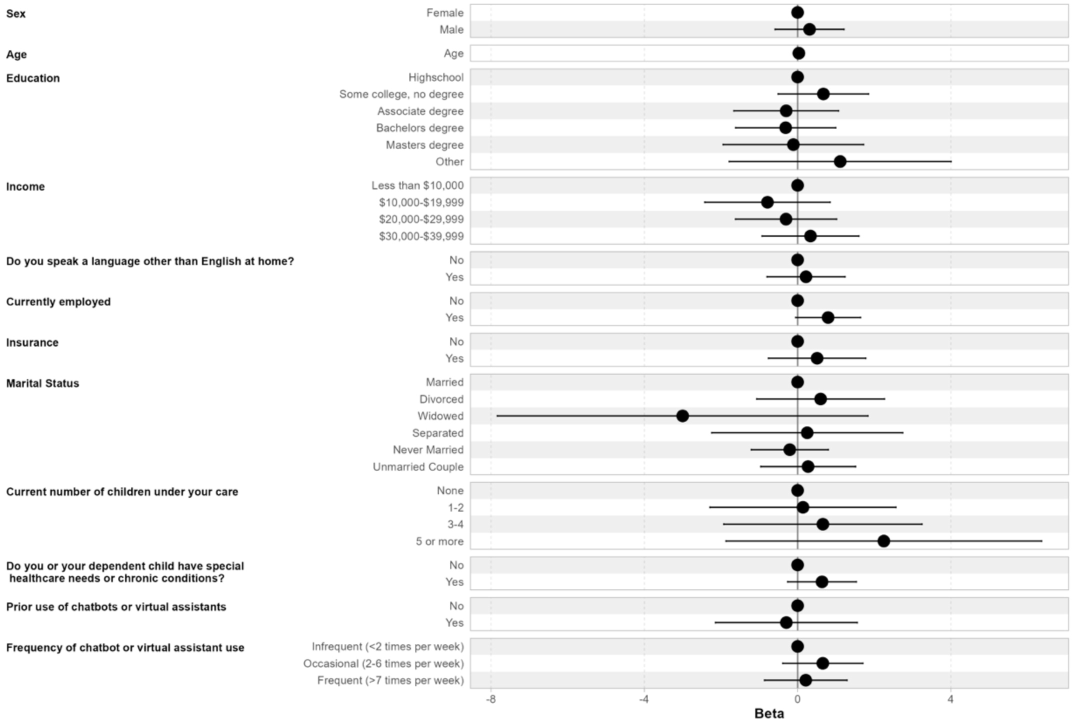

### Figure S8.5 HRSN and Usability Forest Plot

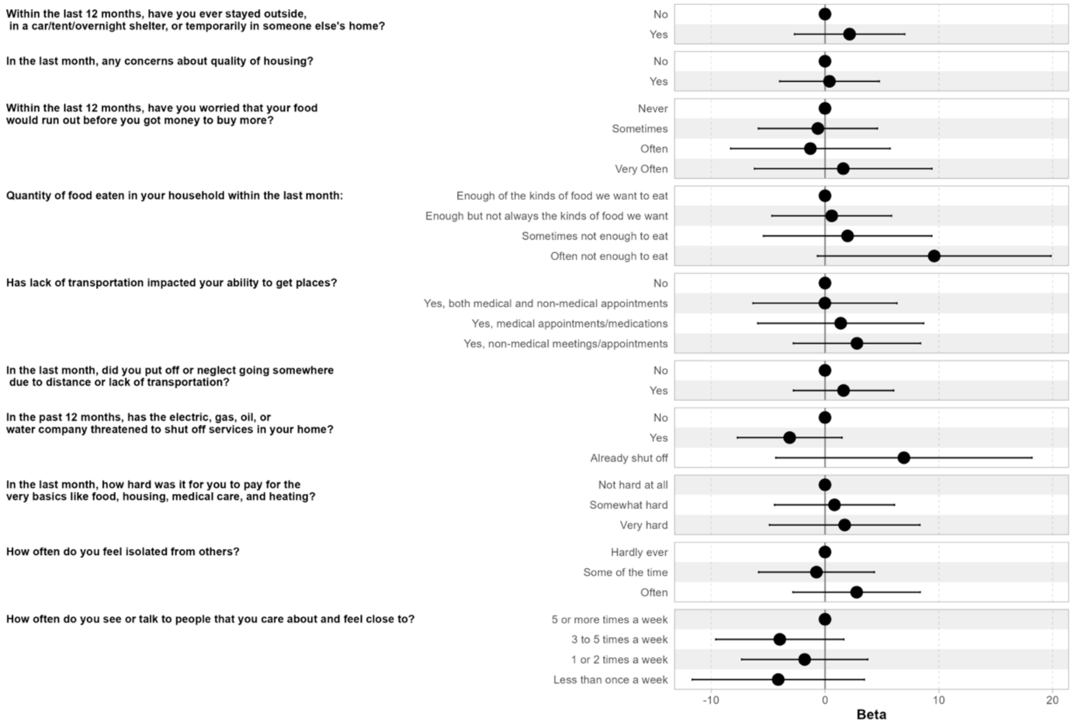


### Figure S8.6 HRSN and Trust Forest Plot

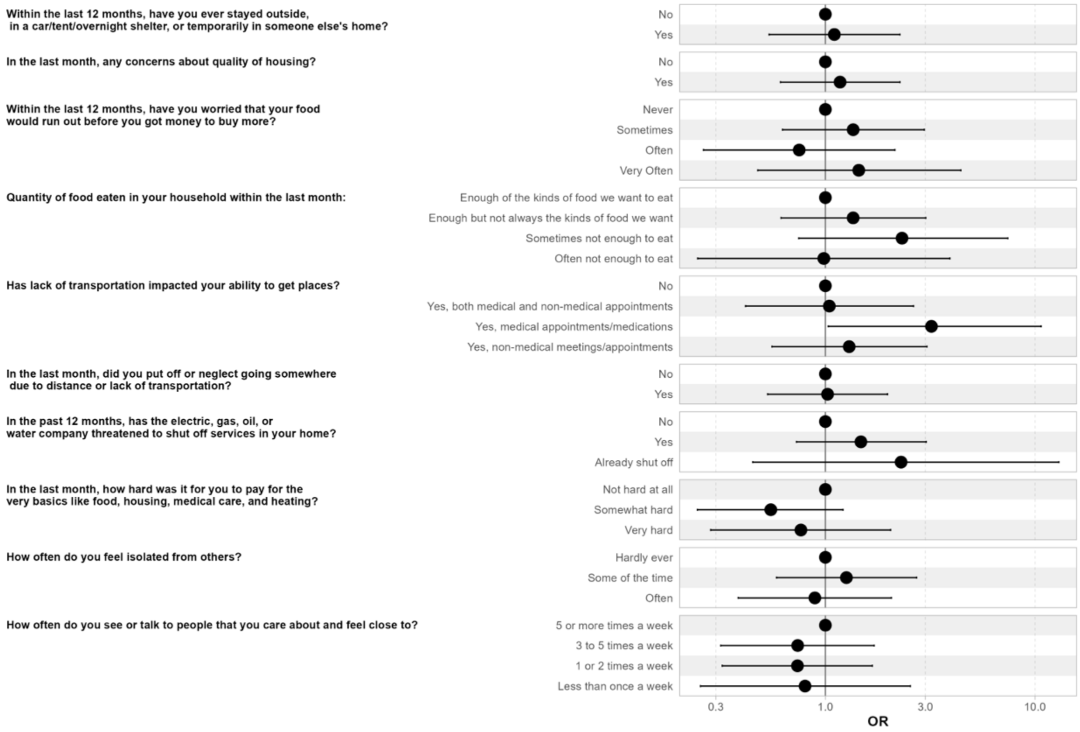

### Figure S8.7 HRSN and Task Load Forest Plot

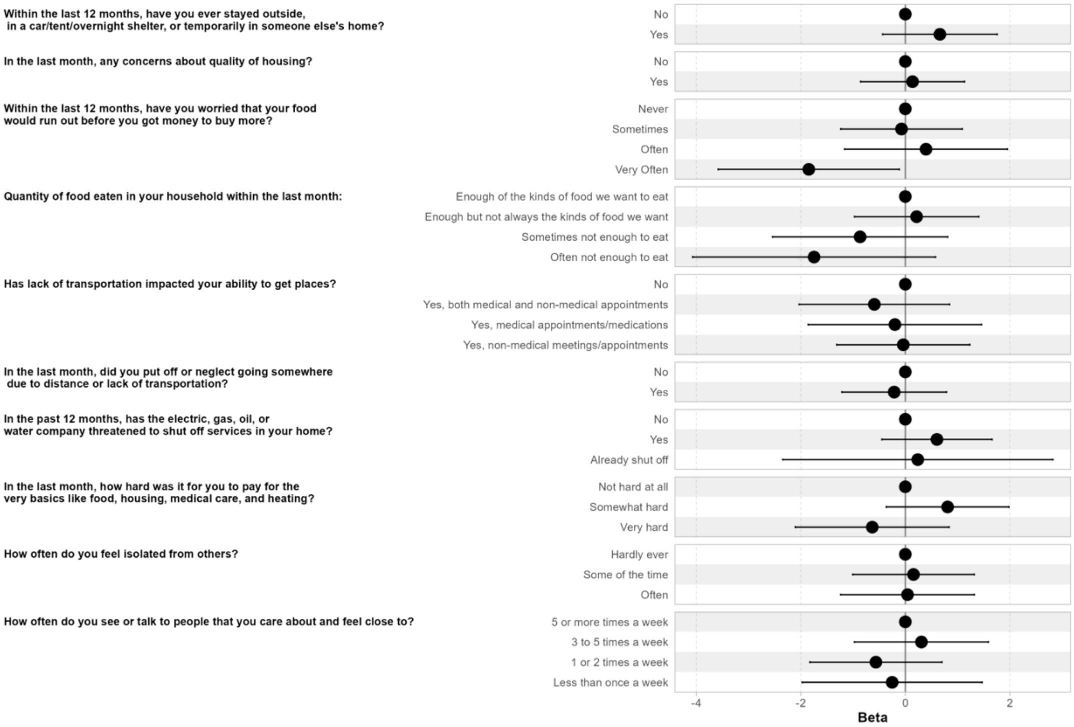


### Figure S8.8 HRSN and Satisfaction Forest Plot

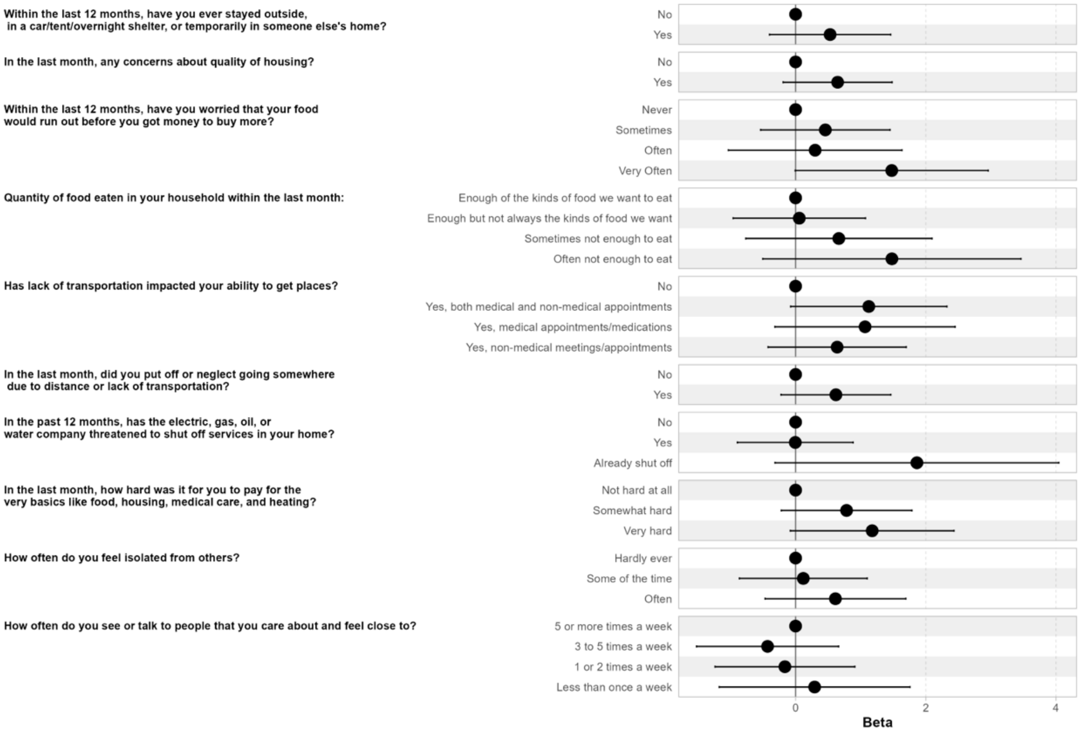

### Supplementary Appendix 9

#### Table S9.1 Education and Usability Pairwise Comparison

| contrast | estimate | SE | lower.CL | upper.CL |
|---|---|---|---|---|
| Some college, no degree - Highschool | 1.02 | 3.06 | -7.85 | 9.89 |
| Associate degree - Highschool | -3.72 | 3.55 | -14.00 | 6.56 |
| Associate degree - Some college, no degree | -4.74 | 3.27 | -14.19 | 4.72 |
| Bachelors degree - Highschool | -5.91 | 3.40 | -15.74 | 3.93 |
| Bachelors degree - Some college, no degree | -6.92 | 3.10 | -15.90 | 2.05 |
| Bachelors degree - Associate degree | -2.19 | 3.58 | -12.56 | 8.18 |
| Masters degree - Highschool | -4.89 | 4.76 | -18.66 | 8.89 |
| Masters degree - Some college, no degree | -5.91 | 4.55 | -19.08 | 7.27 |
| Masters degree - Associate degree | -1.17 | 4.89 | -15.33 | 12.99 |
| Masters degree - Bachelors degree | 1.02 | 4.78 | -12.82 | 14.86 |

#### Table S9.2 Income and Usability Pairwise Comparison

| contrast | estimate | SE | lower.CL | upper.CL |
|---|---|---|---|---|
| ($10,000-$15,999) - Less than $10,000 | -8.46 | 7.75 | -29.91 | 12.99 |
| ($16,000-$19,999) - Less than $10,000 | -4.12 | 4.60 | -16.85 | 8.61 |
| ($16,000-$19,999) - ($10,000-$15,999) | 4.33 | 8.05 | -17.95 | 26.62 |
| ($20,000-$29,999) - Less than $10,000 | -2.24 | 3.47 | -11.86 | 7.38 |
| ($20,000-$29,999) - ($10,000-$15,999) | 6.22 | 7.46 | -14.45 | 26.88 |
| ($20,000-$29,999) - ($16,000-$19,999) | 1.88 | 4.10 | -9.48 | 13.25 |
| ($30,000-$39,999) - Less than $10,000 | 0.40 | 3.33 | -8.81 | 9.60 |
| ($30,000-$39,999) - ($10,000-$15,999) | 8.85 | 7.40 | -11.63 | 29.33 |
| ($30,000-$39,999) - ($16,000-$19,999) | 4.52 | 3.98 | -6.50 | 15.54 |
| ($30,000-$39,999) - ($20,000-$29,999) | 2.64 | 2.60 | -4.57 | 9.84 |

#### Table S9.3 Children and Usability Pairwise Comparison

| contrast | estimate | SE | lower.CL | upper.CL |
|---|---|---|---|---|
| (1-2) - None | 6.58 | 6.35 | -9.96 | 23.12 |
| (3-4) - None | 7.45 | 6.77 | -10.17 | 25.08 |
| (3-4) - (1-2) | 0.87 | 2.93 | -6.76 | 8.51 |
| 5 or more - None | 16.00 | 10.78 | -12.08 | 44.08 |
| 5 or more - (1-2) | 9.42 | 8.89 | -13.74 | 32.58 |
| 5 or more - (3-4) | 8.55 | 9.20 | -15.40 | 32.50 |

#### Table S9.4 Education and Task Load Pairwise Comparison

| contrast | estimate | SE | lower.CL | upper.CL |
|---|---|---|---|---|
| Some college, no degree - Highschool | -1.27 | 0.68 | -3.22 | 0.69 |
| Associate degree - Highschool | -0.32 | 0.78 | -2.59 | 1.95 |
| Associate degree - Some college, no degree | 0.94 | 0.72 | -1.15 | 3.03 |
| Bachelors degree - Highschool | 0.89 | 0.75 | -1.28 | 3.06 |
| Bachelors degree - Some college, no degree | 2.15 | 0.68 | 0.17 | 4.13 |
| Bachelors degree - Associate degree | 1.21 | 0.79 | -1.08 | 3.50 |
| Masters degree - Highschool | 1.91 | 1.05 | -1.13 | 4.95 |
| Masters degree - Some college, no degree | 3.17 | 1.00 | 0.27 | 6.08 |
| Masters degree - Associate degree | 2.23 | 1.08 | -0.89 | 5.36 |
| Masters degree - Bachelors degree | 1.02 | 1.05 | -2.04 | 4.07 |

#### Table S9.5 Income and Task Load Pairwise Comparison

| contrast | estimate | SE | lower.CL | upper.CL |
|---|---|---|---|---|
| ($10,000-$15,999) - Less than $10,000 | -0.36 | 1.77 | -5.25 | 4.54 |
| ($16,000-$19,999) - Less than $10,000 | 0.74 | 1.05 | -2.17 | 3.64 |
| ($16,000-$19,999) - ($10,000-$15,999) | 1.10 | 1.84 | -3.99 | 6.18 |
| ($20,000-$29,999) - Less than $10,000 | 0.97 | 0.79 | -1.23 | 3.16 |
| ($20,000-$29,999) - ($10,000-$15,999) | 1.33 | 1.70 | -3.39 | 6.04 |
| ($20,000-$29,999) - ($16,000-$19,999) | 0.23 | 0.94 | -2.37 | 2.82 |
| ($30,000-$39,999) - Less than $10,000 | 0.92 | 0.76 | -1.18 | 3.03 |
| ($30,000-$39,999) - ($10,000-$15,999) | 1.28 | 1.69 | -3.39 | 5.96 |
| ($30,000-$39,999) - ($16,000-$19,999) | 0.19 | 0.91 | -2.33 | 2.70 |
| ($30,000-$39,999) - ($20,000-$29,999) | -0.04 | 0.59 | -1.68 | 1.60 |

#### Table S9.6 Children and Task Load Pairwise Comparison

| contrast | estimate | SE | lower.CL | upper.CL |
|---|---|---|---|---|
| (1-2) - None | -1.38 | 1.45 | -5.16 | 2.40 |
| (3-4) - None | -1.70 | 1.55 | -5.73 | 2.33 |
| (3-4) - (1-2) | -0.32 | 0.67 | -2.07 | 1.42 |
| 5 or more - None | -1.33 | 2.46 | -7.75 | 5.08 |
| 5 or more - (1-2) | 0.04 | 2.03 | -5.25 | 5.34 |
| 5 or more - (3-4) | 0.36 | 2.10 | -5.11 | 5.84 |

#### Table S9.7 Education and Satisfaction Pairwise Comparison

| contrast | estimate | SE | lower.CL | upper.CL |
|---|---|---|---|---|
| Some college, no degree - Highschool | 0.67 | 0.60 | -1.06 | 2.40 |
| Associate degree - Highschool | -0.30 | 0.69 | -2.30 | 1.71 |
| Associate degree - Some college, no degree | -0.97 | 0.64 | -2.82 | 0.88 |
| Bachelors degree - Highschool | -0.31 | 0.66 | -2.23 | 1.61 |
| Bachelors degree - Some college, no degree | -0.98 | 0.60 | -2.74 | 0.77 |
| Bachelors degree - Associate degree | -0.01 | 0.70 | -2.04 | 2.01 |
| Masters degree - Highschool | -0.11 | 0.93 | -2.80 | 2.58 |
| Masters degree - Some college, no degree | -0.78 | 0.89 | -3.35 | 1.79 |
| Masters degree - Associate degree | 0.19 | 0.95 | -2.58 | 2.95 |
| Masters degree - Bachelors degree | 0.20 | 0.93 | -2.50 | 2.90 |

#### Table S9.8 Income and Satisfaction Pairwise Comparison

| contrast | estimate | SE | lower.CL | upper.CL |
|---|---|---|---|---|
| ($10,000-$15,999) - Less than $10,000 | -3.05 | 1.47 | -7.13 | 1.02 |
| ($16,000-$19,999) - Less than $10,000 | -0.22 | 0.87 | -2.64 | 2.20 |
| ($16,000-$19,999) - ($10,000-$15,999) | 2.83 | 1.53 | -1.40 | 7.07 |
| ($20,000-$29,999) - Less than $10,000 | -0.30 | 0.66 | -2.13 | 1.53 |
| ($20,000-$29,999) - ($10,000-$15,999) | 2.75 | 1.42 | -1.18 | 6.68 |
| ($20,000-$29,999) - ($16,000-$19,999) | -0.08 | 0.78 | -2.24 | 2.08 |
| ($30,000-$39,999) - Less than $10,000 | 0.34 | 0.63 | -1.41 | 2.09 |
| ($30,000-$39,999) - ($10,000-$15,999) | 3.39 | 1.41 | -0.50 | 7.28 |
| ($30,000-$39,999) - ($16,000-$19,999) | 0.56 | 0.76 | -1.54 | 2.65 |
| ($30,000-$39,999) - ($20,000-$29,999) | 0.64 | 0.49 | -0.73 | 2.01 |

#### Table S9.9 Children and Satisfaction Pairwise Comparison

| contrast | estimate | SE | lower.CL | upper.CL |
|---|---|---|---|---|
| (1-2) - None | 0.14 | 1.23 | -3.05 | 3.33 |
| (3-4) - None | 0.66 | 1.31 | -2.74 | 4.06 |
| (3-4) - (1-2) | 0.52 | 0.57 | -0.95 | 1.99 |
| 5 or more - None | 2.25 | 2.08 | -3.17 | 7.67 |
| 5 or more - (1-2) | 2.11 | 1.72 | -2.36 | 6.58 |
| 5 or more - (3-4) | 1.59 | 1.77 | -3.03 | 6.21 |

#### Table S9.10 Food Quantity and Usability Pairwise Comparison

| contrast | estimate | SE | lower. CL | upper. CL |
|---|---|---|---|---|
| Enough but not always the kinds of food we want - Enough of the kinds of food we want to eat | 0.59 | 2.65 | -6.33 | 7.50 |
| Sometimes not enough to eat - Enough of the kinds of food we want to eat | 1.98 | 3.74 | -7.75 | 11.71 |
| Sometimes not enough to eat - Enough but not always the kinds of food we want | 1.39 | 3.34 | -7.29 | 10.08 |
| Often not enough to eat - Enough of the kinds of food we want to eat | 9.59 | 5.18 | -3.90 | 23.09 |
| Often not enough to eat - Enough but not always the kinds of food we want | 9.01 | 4.90 | -3.76 | 21.77 |
| Often not enough to eat - Sometimes not enough to eat | 7.61 | 5.56 | -6.87 | 22.10 |

#### Table S9.11 Utility Shutoff and Usability Pairwise Comparison

| contrast | estimate | SE | lower.CL | upper.CL |
|---|---|---|---|---|
| Yes - No | -3.10 | 2.32 | -8.61 | 2.40 |
| Already shut off - No | 6.93 | 5.69 | -6.56 | 20.42 |
| Already shut off - Yes | 10.04 | 5.82 | -3.77 | 23.84 |

#### Table S9.12 Needs Affordability and Usability Pairwise Comparison

| contrast | estimate | SE | lower.CL | upper.CL |
|---|---|---|---|---|
| Somewhat hard - Not hard at all | 0.83 | 2.66 | -5.49 | 7.15 |
| Very hard - Not hard at all | 1.72 | 3.34 | -6.19 | 9.64 |
| Very hard - Somewhat hard | 0.90 | 2.91 | -6.00 | 7.79 |

#### Table S9.13 Social Isolation and Usability Pairwise Comparison

| contrast | estimate | SE | lower.CL | upper.CL |
|---|---|---|---|---|
| 5 or more times a week - 3 to 5 times a week | 3.98 | 2.84 | -3.43 | 11.38 |
| 5 or more times a week - 1 or 2 times a week | 1.79 | 2.80 | -5.49 | 9.08 |
| 5 or more times a week - Less than once a week | 4.11 | 3.82 | -5.84 | 14.06 |
| 3 to 5 times a week - 1 or 2 times a week | -2.18 | 2.98 | -9.94 | 5.57 |
| 3 to 5 times a week - Less than once a week | 0.13 | 3.95 | -10.16 | 10.43 |
| 1 or 2 times a week - Less than once a week | 2.32 | 3.92 | -7.89 | 12.53 |

#### Table S9.14 Food Quantity and Task Load Pairwise Comparison

| contrast | estimate | SE | lower.CL | upper.CL |
|---|---|---|---|---|
| Enough but not always the kinds of food we want - Enough of the kinds of food we want to eat | 0.22 | 0.60 | -1.35 | 1.78 |
| Sometimes not enough to eat - Enough of the kinds of food we want to eat | -0.87 | 0.85 | -3.07 | 1.34 |
| Sometimes not enough to eat - Enough but not always the kinds of food we want | -1.08 | 0.76 | -3.05 | 0.89 |
| Often not enough to eat - Enough of the kinds of food we want to eat | -1.75 | 1.17 | -4.81 | 1.31 |
| Often not enough to eat - Enough but not always the kinds of food we want | -1.96 | 1.11 | -4.86 | 0.93 |
| Often not enough to eat - Sometimes not enough to eat | -0.88 | 1.26 | -4.16 | 2.40 |

#### Table S9.15 Utility Shutoff and Task Load Pairwise Comparison

| contrast | estimate | SE | lower.CL | upper.CL |
|---|---|---|---|---|
| Yes - No | 0.61 | 0.53 | -0.66 | 1.87 |
| Already shut off - No | 0.24 | 1.31 | -2.86 | 3.34 |
| Already shut off - Yes | -0.37 | 1.34 | -3.54 | 2.81 |

#### Table S9.16 Needs Affordability and Task Load Pairwise Comparison

| contrast | estimate | SE | lower.CL | upper.CL |
|---|---|---|---|---|
| Somewhat hard - Not hard at all | 0.81 | 0.59 | -0.60 | 2.22 |
| Very hard - Not hard at all | -0.63 | 0.74 | -2.40 | 1.13 |
| Very hard - Somewhat hard | -1.44 | 0.65 | -2.98 | 0.09 |

#### Table S9.17 Social Isolation and Task Load Pairwise Comparison

| contrast | estimate | SE | lower.CL | upper.CL |
|---|---|---|---|---|
| 5 or more times a week - 3 to 5 times a week | -0.31 | 0.65 | -2.00 | 1.38 |
| 5 or more times a week - 1 or 2 times a week | 0.56 | 0.64 | -1.10 | 2.23 |
| 5 or more times a week - Less than once a week | 0.25 | 0.87 | -2.02 | 2.52 |
| 3 to 5 times a week - 1 or 2 times a week | 0.87 | 0.68 | -0.90 | 2.64 |
| 3 to 5 times a week - Less than once a week | 0.56 | 0.90 | -1.79 | 2.91 |
| 1 or 2 times a week - Less than once a week | -0.31 | 0.89 | -2.64 | 2.02 |

#### Table S9.18 Food Quantity and Satisfaction Pairwise Comparison

| contrast | estimate | SE | lower.CL | upper.CL |
|---|---|---|---|---|
| Enough but not always the kinds of food we want - Enough of the kinds of food we want to eat | 0.06 | 0.51 | -1.28 | 1.39 |
| Sometimes not enough to eat - Enough of the kinds of food we want to eat | 0.66 | 0.72 | -1.22 | 2.55 |
| Sometimes not enough to eat - Enough but not always the kinds of food we want | 0.61 | 0.64 | -1.07 | 2.29 |
| Often not enough to eat - Enough of the kinds of food we want to eat | 1.48 | 1.00 | -1.13 | 4.09 |
| Often not enough to eat - Enough but not always the kinds of food we want | 1.42 | 0.95 | -1.04 | 3.89 |
| Often not enough to eat - Sometimes not enough to eat | 0.82 | 1.08 | -1.99 | 3.62 |

#### Table S9.19 Utility Shutoff and Satisfaction Pairwise Comparison

| contrast | estimate | SE | lower.CL | upper.CL |
|---|---|---|---|---|
| Yes - No | -0.00 | 0.45 | -1.07 | 1.06 |
| Already shut off - No | 1.86 | 1.10 | -0.75 | 4.47 |
| Already shut off - Yes | 1.87 | 1.13 | -0.80 | 4.54 |

#### Table S9.20 Needs Affordability and Satisfaction Pairwise Comparison

| contrast | estimate | SE | lower.CL | upper.CL |
|---|---|---|---|---|
| Somewhat hard - Not hard at all | 0.78 | 0.51 | -0.42 | 1.98 |
| Very hard - Not hard at all | 1.18 | 0.63 | -0.33 | 2.68 |
| Very hard - Somewhat hard | 0.39 | 0.55 | -0.92 | 1.71 |

#### Table S9.20 Social Isolation and Satisfaction Pairwise Comparison

| contrast | estimate | SE | lower.CL | upper.CL |
|---|---|---|---|---|
| 5 or more times a week - 3 to 5 times a week | 0.43 | 0.55 | -1.01 | 1.87 |
| 5 or more times a week - 1 or 2 times a week | 0.16 | 0.54 | -1.25 | 1.58 |
| 5 or more times a week - Less than once a week | -0.29 | 0.74 | -2.22 | 1.64 |
| 3 to 5 times a week - 1 or 2 times a week | -0.27 | 0.58 | -1.77 | 1.24 |
| 3 to 5 times a week - Less than once a week | -0.72 | 0.77 | -2.72 | 1.27 |
| 1 or 2 times a week - Less than once a week | -0.46 | 0.76 | -2.44 | 1.52 |

### Supplementary Appendix 10

#### Table S10.1 Education Mean Trust Class Estimate

| education | mean.class | SE | asymp.LCL | asymp.UCL |
|---|---|---|---|---|
| Highschool | 2.99 | 0.16 | 2.68 | 3.29 |
| Some college, no degree | 3.24 | 0.11 | 3.02 | 3.46 |
| Associate degree | 3.01 | 0.16 | 2.70 | 3.32 |
| Bachelors degree | 2.94 | 0.16 | 2.63 | 3.25 |
| Masters degree | 3.06 | 0.27 | 2.54 | 3.58 |
| None of the above | 2.04 | 0.70 | 0.68 | 3.41 |

#### Table S10.2 Income Mean Trust Class Estimate

| income | mean.class | SE | asymp.LCL | asymp.UCL |
|---|---|---|---|---|
| Less than $10,000 | 2.90 | 0.19 | 2.53 | 3.26 |
| $10,000-$15,999 | 2.02 | 0.48 | 1.08 | 2.95 |
| $16,000-$19,999 | 2.97 | 0.22 | 2.54 | 3.41 |
| $20,000-$29,999 | 3.19 | 0.12 | 2.95 | 3.42 |
| $30,000-$39,999 | 3.12 | 0.10 | 2.93 | 3.32 |

#### Table S10.3 Kids Mean Trust Class Estimate

| kids | mean.class | SE | asymp.LCL | asymp.UCL |
|---|---|---|---|---|
| None | 2.49 | 0.38 | 1.73 | 3.24 |
| 1-2 | 3.08 | 0.08 | 2.92 | 3.23 |
| 3-4 | 3.14 | 0.17 | 2.82 | 3.46 |
| 5 or more | 3.44 | 0.45 | 2.56 | 4.32 |

#### Table S10.4 Food Quantity Mean Trust Class Estimate

| contrast | estimate | SE | lower.CL | upper.CL |
|---|---|---|---|---|
| Enough but not always the kinds of food we want - Enough of the kinds of food we want to eat | 0.22 | 0.60 | -1.35 | 1.78 |
| Sometimes not enough to eat - Enough of the kinds of food we want to eat | -0.87 | 0.85 | -3.07 | 1.34 |
| Sometimes not enough to eat - Enough but not always the kinds of food we want | -1.08 | 0.76 | -3.05 | 0.89 |
| Often not enough to eat - Enough of the kinds of food we want to eat | -1.75 | 1.17 | -4.81 | 1.31 |

| | | | | |
|---|---|---|---|---|
| Often not enough to eat - Enough but not always the kinds of food we want | -1.96 | 1.11 | -4.86 | 0.93 |
| Often not enough to eat - Sometimes not enough to eat | -0.88 | 1.26 | -4.16 | 2.40 |

#### Table S10.5 Utility Shutoff Mean Trust Class Estimate

| contrast | estimate | SE | lower.CL | upper.CL |
|---|---|---|---|---|
| Yes - No | 0.61 | 0.53 | -0.66 | 1.87 |
| Already shut off - No | 0.24 | 1.31 | -2.86 | 3.34 |
| Already shut off - Yes | -0.37 | 1.34 | -3.54 | 2.81 |

#### Table S10.6 Needs Affordability Mean Trust Class Estimate

| contrast | estimate | SE | lower.CL | upper.CL |
|---|---|---|---|---|
| Somewhat hard - Not hard at all | 0.81 | 0.59 | -0.60 | 2.22 |
| Very hard - Not hard at all | -0.63 | 0.74 | -2.40 | 1.13 |
| Very hard - Somewhat hard | -1.44 | 0.65 | -2.98 | 0.09 |

#### Table S10.7 Social Isolation Mean Trust Class Estimate

| contrast | estimate | SE | lower.CL | upper.CL |
|---|---|---|---|---|
| 5 or more times a week - 3 to 5 times a week | -0.31 | 0.65 | -2.00 | 1.38 |
| 5 or more times a week - 1 or 2 times a week | 0.56 | 0.64 | -1.10 | 2.23 |
| 5 or more times a week - Less than once a week | 0.25 | 0.87 | -2.02 | 2.52 |
| 3 to 5 times a week - 1 or 2 times a week | 0.87 | 0.68 | -0.90 | 2.64 |
| 3 to 5 times a week - Less than once a week | 0.56 | 0.90 | -1.79 | 2.91 |
| 1 or 2 times a week - Less than once a week | -0.31 | 0.89 | -2.64 | 2.02 |
