## Supplementary material for "Evaluating User Experiences with an AI Chatbot for Health-Related Social Needs: A Cross-Sectional Mixed Methods Study": STROBE

STROBE Statement—checklist of items that should be included in reports of observational studies

| | Item No. | Recommendation | Page No. | Relevant text from manuscript |
|---|---|---|---|---|
| **Title and abstract** | 1 | (*a*) Indicate the study's design with a commonly used term in the title or the abstract | 1 | *Cross-Sectional Mixed-Methods* |
| | | (*b*) Provide in the abstract an informative and balanced summary of what was done and what was found | 1 | *Background: Health-related…* |
| **Introduction** | | | | |
| Background/rationale | 2 | Explain the scientific background and rationale for the investigation being reported | 3 | *Health-related social needs…* |
| Objectives | 3 | State specific objectives, including any prespecified hypotheses | 3 | *…our findings aim to inform…* |
| **Methods** | | | | |
| Study design | 4 | Present key elements of study design early in the paper | 4 | *…conducted a cross-sectional…* |
| Setting | 5 | Describe the setting, locations, and relevant dates, including periods of recruitment, exposure, follow-up, and data collection | 6 | *…an online research platform…* |
| Participants | 6 | (*a*) *Cohort study*—Give the eligibility criteria, and the sources and methods of selection of participants. Describe methods of follow-up<br>*Case-control study*—Give the eligibility criteria, and the sources and methods of case ascertainment and control selection. Give the rationale for the choice of cases and controls<br>***Cross-sectional study***—Give the eligibility criteria, and the sources and methods of selection of participants | 6 | *Inclusion criteria required…* |
| | | (*b*) *Cohort study*—For matched studies, give matching criteria and number of exposed and unexposed<br>*Case-control study*—For matched studies, give matching criteria and the number of controls per case | NA | *NA* |
| Variables | 7 | Clearly define all outcomes, exposures, predictors, potential confounders, and effect modifiers. Give diagnostic criteria, if applicable | 7 | *Quantitative data collection…* |
| Data sources/ measurement | 8* | For each variable of interest, give sources of data and details of methods of assessment (measurement). Describe comparability of assessment methods if there is more than one group | 7 | *…HRSNs were adapted from…* |
| Bias | 9 | Describe any efforts to address potential sources of bias | 8 | *Codes were refined iteratively…* |
| Study size | 10 | Explain how the study size was arrived at | 6 | *Recruitment ended after…* |

Continued on next page

| | | | | |
|---|---|---|---|---|
| Quantitative variables | 11 | Explain how quantitative variables were handled in the analyses. If applicable, describe which groupings were chosen and why | 7 | *As exploratory measures, the…* |
| Statistical methods | 12 | (*a*) Describe all statistical methods, including those used to control for confounding | 7 | *…were modelled via linear…* |
| | | (*b*) Describe any methods used to examine subgroups and interactions | 7 | *…a pairwise analysis between…* |
| | | (*c*) Explain how missing data were addressed | 9 | *…were included in the analysis.* |
| | | (*d*) *Cohort study*—If applicable, explain how loss to follow-up was addressed<br>*Case-control study*—If applicable, explain how matching of cases and controls was addressed<br>***Cross-sectional study***—If applicable, describe analytical methods taking account of sampling strategy | 7 | *…category-wise regressions…* |
| | | (*e*) Describe any sensitivity analyses | 7 | *…to quantify suggestive trend...* |
| **Results** | | | | |
| Participants | 13* | (a) Report numbers of individuals at each stage of study—eg numbers potentially eligible, examined for eligibility, confirmed eligible, included in the study, completing follow-up, and analysed | 9 | *We recruited 167 participants…* |
| | | (b) Give reasons for non-participation at each stage | 9 | *76.6% completed all activities…* |
| | | (c) Consider use of a flow diagram | 7 | ***Figure 2.*** *Study design…* |
| Descriptive data | 14* | (a) Give characteristics of study participants (eg demographic, clinical, social) and information on exposures and potential confounders | 9 | *The sample (N=128) was…* |
| | | (b) Indicate number of participants with missing data for each variable of interest | 9 | ***Table 1.*** *Demographics of...* |
| | | (c) *Cohort study*—Summarise follow-up time (eg, average and total amount) | NA | *NA* |
| Outcome data | 15* | *Cohort study*—Report numbers of outcome events or summary measures over time | NA | *NA* |
| | | *Case-control study*—Report numbers in each exposure category, or summary measures of exposure | NA | *NA* |
| | | ***Cross-sectional study***—Report numbers of outcome events or summary measures | 12 | *…rated the chatbot as being…* |
| Main results | 16 | (*a*) Give unadjusted estimates and, if applicable, confounder-adjusted estimates and their precision (eg, 95% confidence interval). Make clear which confounders were adjusted for and why they were included | 12 | *we highlight response options…* |
| | | (*b*) Report category boundaries when continuous variables were categorized | 12 | *…as income and education.* |
| | | (*c*) If relevant, consider translating estimates of relative risk into absolute risk for a meaningful time-period | 11 | *…grouped by short-term…* |

Continued on next page

| | | | | |
|---|---|---|---|---|
| Other analyses | 17 | Report other analyses done—eg analyses of subgroups and interactions, and sensitivity analyses | 15 | *To better understand user…* |
| **Discussion** | | | | |
| Key results | 18 | Summarise key results with reference to study objectives | 17 | *The present mixed-methods…* |
| Limitations | 19 | Discuss limitations of the study, taking into account sources of potential bias or imprecision. Discuss both direction and magnitude of any potential bias | 19 | *…several limitations warrant…* |
| Interpretation | 20 | Give a cautious overall interpretation of results considering objectives, limitations, multiplicity of analyses, results from similar studies, and other relevant evidence | 17 | *The study sample was largely…* |
| Generalisability | 21 | Discuss the generalisability (external validity) of the study results | 20 | *…setting limit generalizability.* |
| **Other information** | | | | |
| Funding | 22 | Give the source of funding and the role of the funders for the present study and, if applicable, for the original study on which the present article is based | 21 | *This research received "BIIG…* |

*Give information separately for cases and controls in case-control studies and, if applicable, for exposed and unexposed groups in cohort and cross-sectional studies.

**Note:** An Explanation and Elaboration article discusses each checklist item and gives methodological background and published examples of transparent reporting. The STROBE checklist is best used in conjunction with this article (freely available on the Web sites of PLoS Medicine at http://www.plosmedicine.org/, Annals of Internal Medicine at http://www.annals.org/, and Epidemiology at http://www.epidem.com/). Information on the STROBE Initiative is available at www.strobe-statement.org.
